# Dynamic changes in human single cell transcriptional signatures during fatal sepsis

**DOI:** 10.1101/2021.03.01.21252411

**Authors:** Xinru Qiu, Jiang Li, Jeff Bonenfant, Lukasz Jaroszewski, Walter Klein, Adam Godzik, Meera G. Nair

## Abstract

Systemic infections, especially in patients with chronic diseases, result in sepsis: an explosive, uncoordinated immune response that can lead to multisystem organ failure with a high mortality rate. Sepsis survivors and non-survivors oftentimes have similar clinical phenotypes or sepsis biomarker expression upon diagnosis, suggesting that the dynamics of sepsis in the critical early stage may have an impact on these opposite outcomes. To investigate this, we designed a within-subject study of patients with systemic gram-negative bacterial sepsis with surviving and fatal outcomes and performed single-cell transcriptomic analyses of peripheral blood mononuclear cells (PBMC) collected during the critical period between sepsis recognition and 6 hours. We observed that the largest sepsis-induced expression changes over time in surviving versus fatal sepsis were in CD14+ monocytes, including gene signatures previously reported for sepsis outcomes. We further identify changes in the metabolic pathways of both monocytes and platelets, the emergence of erythroid precursors, and T cell exhaustion signatures, with the most extreme differences occurring between the non-sepsis control and the sepsis non-survivor. Our single-cell observations are consistent with trends from public datasets but also reveal specific effects in individual immune cell populations, which change within hours. In conclusion, this pilot study provides the first single-cell results with a repeated measures design in sepsis to analyze the temporal changes in the immune cell population behavior in surviving or fatal sepsis. These findings indicate that tracking temporal expression changes in specific cell-types could lead to more accurate predictions of sepsis outcomes. We also identify molecular pathways that could be therapeutically controlled to improve the sepsis trajectory toward better outcomes.

**Summary sentence:** Single cell transcriptomics of peripheral blood mononuclear cells in surviving and fatal sepsis reveal inflammatory and metabolic pathways that change within hours of sepsis recognition.

## Introduction

Sepsis is an inflammatory syndrome caused by a systemic infection that can lead to multi-system organ failure and death. Sepsis is responsible for a significant percentage of in-hospital healthcare both in the US and worldwide and is associated with a high mortality rate ^1, 2^. Despite many efforts, no targeted therapeutic agents against sepsis have been developed to assist in the treatment of sepsis. One acknowledged challenge is the complexity of the disease involving the competing interplay between rampant inflammation (cytokine storm) and paradoxically, the almost simultaneous shutdown of the immune system (immunoparalysis) ^3, 4^. Another sepsis paradox is that some patients with nearly identical clinical phenotypes as quantified by qSOFA and APACHE scores, die at every stage of the disease while others survive ^5^. The host response to sepsis has been studied in several blood and PBMC profiling studies with gene-expression or proteomics methods ^6^. These identify several prognostic biomarkers, such as lactate, procalcitonin, C-reactive protein (CRP), ferritin, and erythrocyte sedimentation rate (ESR), which, along with clinical scores, are standardly utilized to evaluate sepsis patients and determine their care ^5^.

However, connecting these high-level observations to accurate clinical outcomes presents an unresolved challenge, likely due to the complexity and heterogeneity of this disease. To gain molecular insight into this heterogeneity, many studies have been conducted to identify a potential sepsis molecular signature, which could aid in diagnosis or treatment ^7^. Recently, the first single-cell analysis of the status of immune cells in sepsis was reported, which identified abnormal monocyte states associated with immune dysregulation ^8^. Here, we apply the same approach to focus on the additional question of immune cell trajectory in sepsis survivor and non-survivor outcomes. We performed single cell transcriptomics analyses in fatal or surviving sepsis using a within-subject study design of peripheral blood mononuclear cells (PBMC) collected from septic patients in the Intensive Care Unit (ICU) at 0 and 6 hours from sepsis recognition. Although a limitation in this study is the small sample size of two sepsis patients, we validated our analyses by comparison with similarly processed PBMC from non-sepsis volunteers and corroborated our findings with available public domain expression datasets. Our timed analyses further reveal the emergence of abnormal immune cells, including new types of cells typical of sepsis but not present in healthy controls, as well as classical cell types present in both sepsis and healthy conditions but with an abnormal gene expression profile. Specifically, we observed that fatal sepsis was associated with expansion of platelets and erythrocyte precursors, and the overall expression of genes related to hypoxic stress and inflammation that were increased over 6 hours, especially in the monocytes. Additionally, the lymphocyte subsets in fatal outcomes expressed genes related to exhaustion. On the other hand, sepsis survival over time involved expression of genes related to recovery from cellular stress, including the regain of cytotoxic effector function for lymphocyte subsets, and an increase in genes related to monocyte migration and chemotaxis. Last, we observed a switch in metabolic state at the cellular level, from oxidative phosphorylation to glycolysis in fatal outcomes. In conclusion, this pilot study, which focused on within-subject analyses of PBMC over time, offers the unique perspective of dynamic immune cell changes in fatal sepsis. Specifically, we identify abnormal immune cell subsets, functional pathways and molecular signatures at the single cell resolution that are associated with fatal or surviving outcomes in sepsis. This study provides foundation data and identifies specific cell subsets and molecular pathways that can be further explored for better prediction of sepsis outcomes.

## Results

### Clinical and immune characteristics of sepsis patients

To gain molecular understanding of the immune state in surviving or non-surviving sepsis outcomes, we performed retrospective single-cell RNA sequencing on PBMC from two septic shock patients. Both patients belonged to a cohort of 45 patients admitted with severe sepsis to the RUHS ICU and enrolled in the study based on Sepsis-3 clinical parameters (manuscript in preparation). Both patients were females, aged between 65 and 70 years old. The first patient expired by 24 hours post-enrollment (NS, non-survivor). The second patient recovered from infection and was discharged from the hospital after 22 days (S, survivor). Both patients suffered from *Escherichia coli* bacteremia and were treated with broad spectrum antibiotics within the first 24 hours. While clinical parameters (qSOFA and APACHE scores) were similar, both patients exhibited different plasma cytokine levels, although in both patients these levels were dramatically elevated compared to baseline non-sepsis volunteers, (Table 1). These results are consistent with the known phenomenon of the sepsis-induced cytokine storm ^4^. Interestingly, re-stimulation of PBMC from the same sepsis patients with lipopolysaccharide led to reduced TNFα secretion as compared to PBMC from non-sepsis controls (Table 1), consistent with monocytic deactivation seen in sepsis immunoparalysis ^9^. Additionally, flow cytometric analysis of PBMC revealed different immune subset distribution with sepsis including reduced monocyte and lymphocyte subsets especially in the non-survivor (Figure 1). We also observed the emergence of cell subsets that we were unable to define with common PBMC surface antibodies (Figure 1C). Together, these data reveal similar and distinct clinical and peripheral immune profiles in sepsis. However, more detailed subsetting of specific immune cells and insights into how temporal changes in their gene expression relate to sepsis outcome were lacking, which we addressed by single cell RNA sequencing.

**Table 1.**
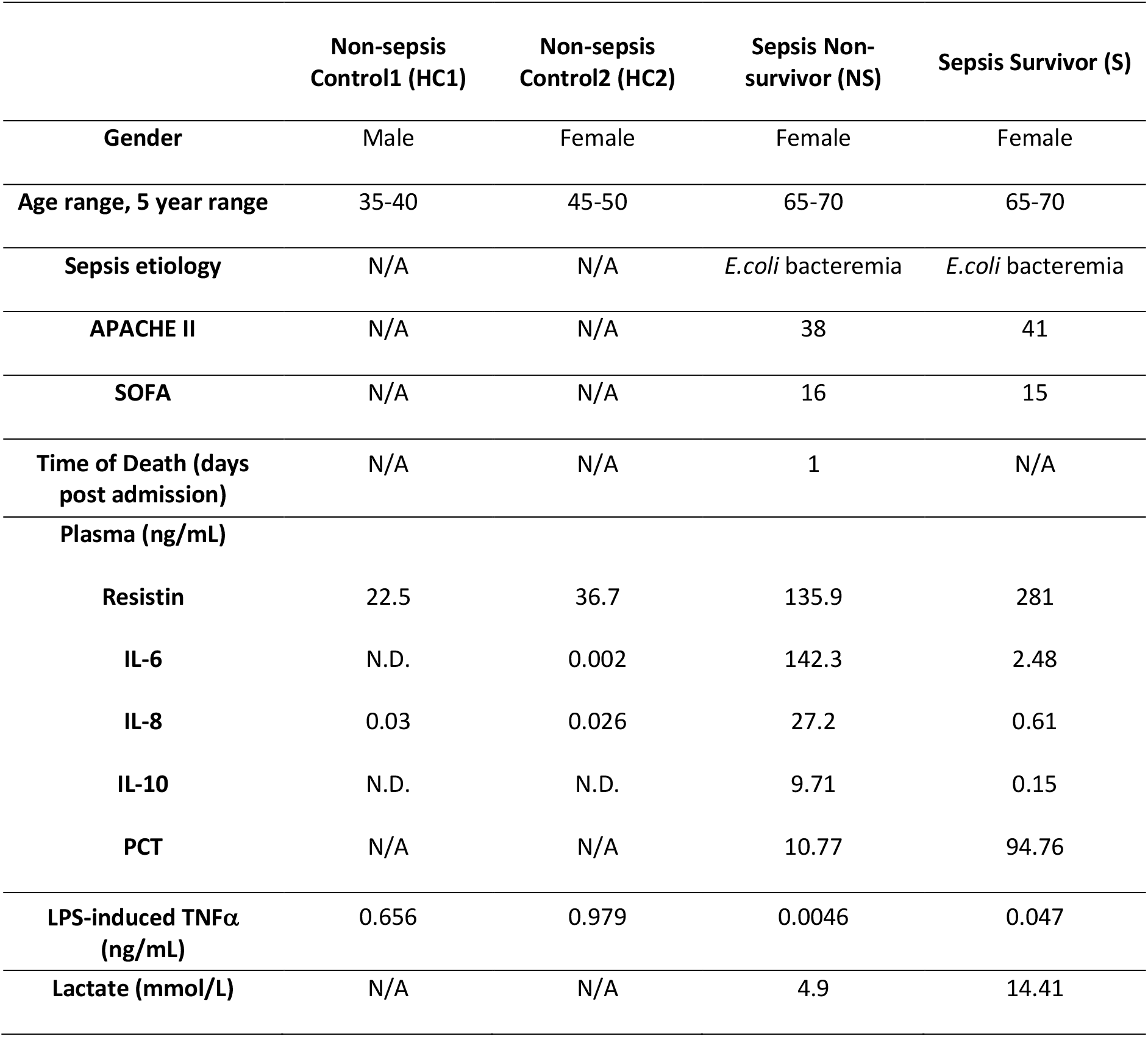
Characteristics of enrolled non-sepsis volunteers and sepsis patients at sepsis recognition (T0). Clinical parameters, cytokine levels in the plasma, and supernatant following LPS stimulation (10 ng/mL) of PBMCs. **APACHE II**: Acute physiology and chronic health evaluation II, **SOFA**: Sequential organ failure assessment, **N.D.:** not detected.

|  | Non-sepsis<br>Control1 (HC1) | Non-sepsis<br>Control2 (HC2) | Sepsis Non-survivor (NS) | Sepsis Survivor (S) |
| --- | --- | --- | --- | --- |
| **Gender** | Male | Female | Female | Female |
| **Age range, 5 year range** | 35-40 | 45-50 | 65-70 | 65-70 |
| **Sepsis etiology** | N/A | N/A | *E.coli* bacteremia | *E.coli* bacteremia |
| **APACHE II** | N/A | N/A | 38 | 41 |
| **SOFA** | N/A | N/A | 16 | 15 |
| **Time of Death (days post admission)** | N/A | N/A | 1 | N/A |
| **Plasma (ng/mL)** |  |  |  |  |
| **Resistin** | 22.5 | 36.7 | 135.9 | 281 |
| **IL-6** | N.D. | 0.002 | 142.3 | 2.48 |
| **IL-8** | 0.03 | 0.026 | 27.2 | 0.61 |
| **IL-10** | N.D. | N.D. | 9.71 | 0.15 |
| **PCT** | N/A | N/A | 10.77 | 94.76 |
| **LPS-induced TNFα (ng/mL)** | 0.656 | 0.979 | 0.0046 | 0.047 |
| **Lactate (mmol/L)** | N/A | N/A | 4.9 | 14.41 |

**Figure 1.**
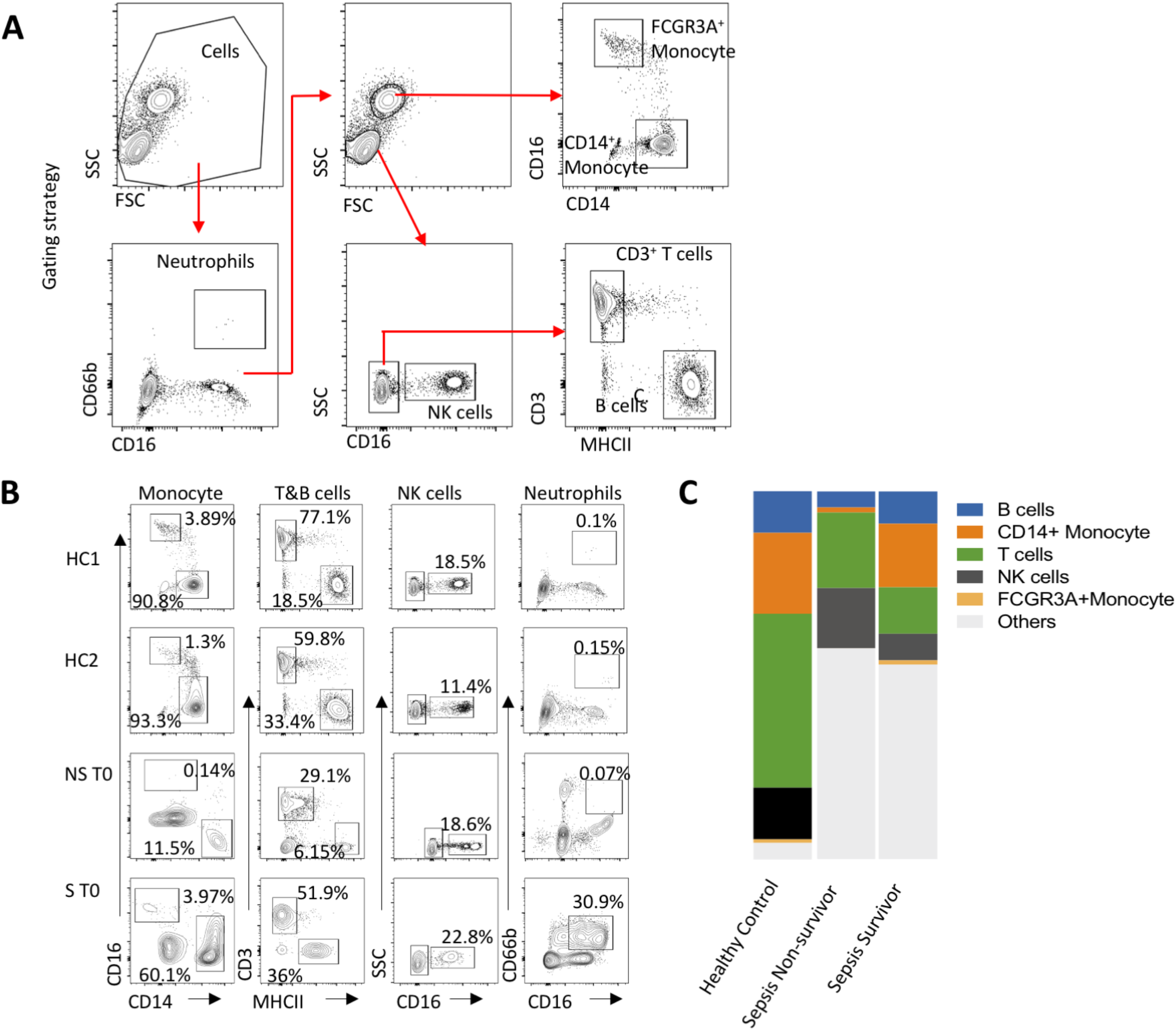
PBMC flow cytometry analysis. Flow cytometric analysis of PBMC from healthy controls (HCs), non-survivor (NS), and survivor (S) sepsis patients at first blood collection (T0). **A)** Gating strategy. **B)** Fractions of immune cell subsets. **C)** Fractions of immune cell types in PBMC.

### Single-cell transcriptomics reveal distinct immune cell subsets in surviving or fatal sepsis

Single-cell transcriptome analysis was performed on a 10X Genomics platform according to a standard pipeline (Figure 2A). After quality filtering (see Materials and methods), the transcriptome profiles of 27,685 cells were collected (15,546 cells from two non-sepsis control samples, 5,758 cells from the two sepsis non-survivor samples, and 6,381 cells from the two sepsis survivor samples). Given that PBMC samples were processed identically, and equivalent numbers of viable cells from each sample were subjected to single cell transcriptomics, the ~3-fold decrease in cell number from sepsis patients that passed quality control is likely caused by the sepsis syndrome and consequent cellular stress and apoptosis. Analysis of the single-cell transcriptomes of all cells from all subjects (Seurat v3 canonical pipeline ^10^) followed by the consensus-based assignment of cell-types to cell clusters (see Materials and Methods section) revealed 10 cell types with the following overall fractions: CD4+ T cells (21.86%), CD8+ T cells (8.73%), B cells (17.75%), Natural killer (NK) cells (9.33%), CD14+ monocytes (22.08%), FCGR3A+ monocytes (6.75%), dendritic cells (DC) (0.06%), erythroid precursor cells (2.45%), platelets (10.66%), and hematopoietic stem and progenitor cells (HSPC) (0.33%) (Figure 2B–D).

**Figure 2.**
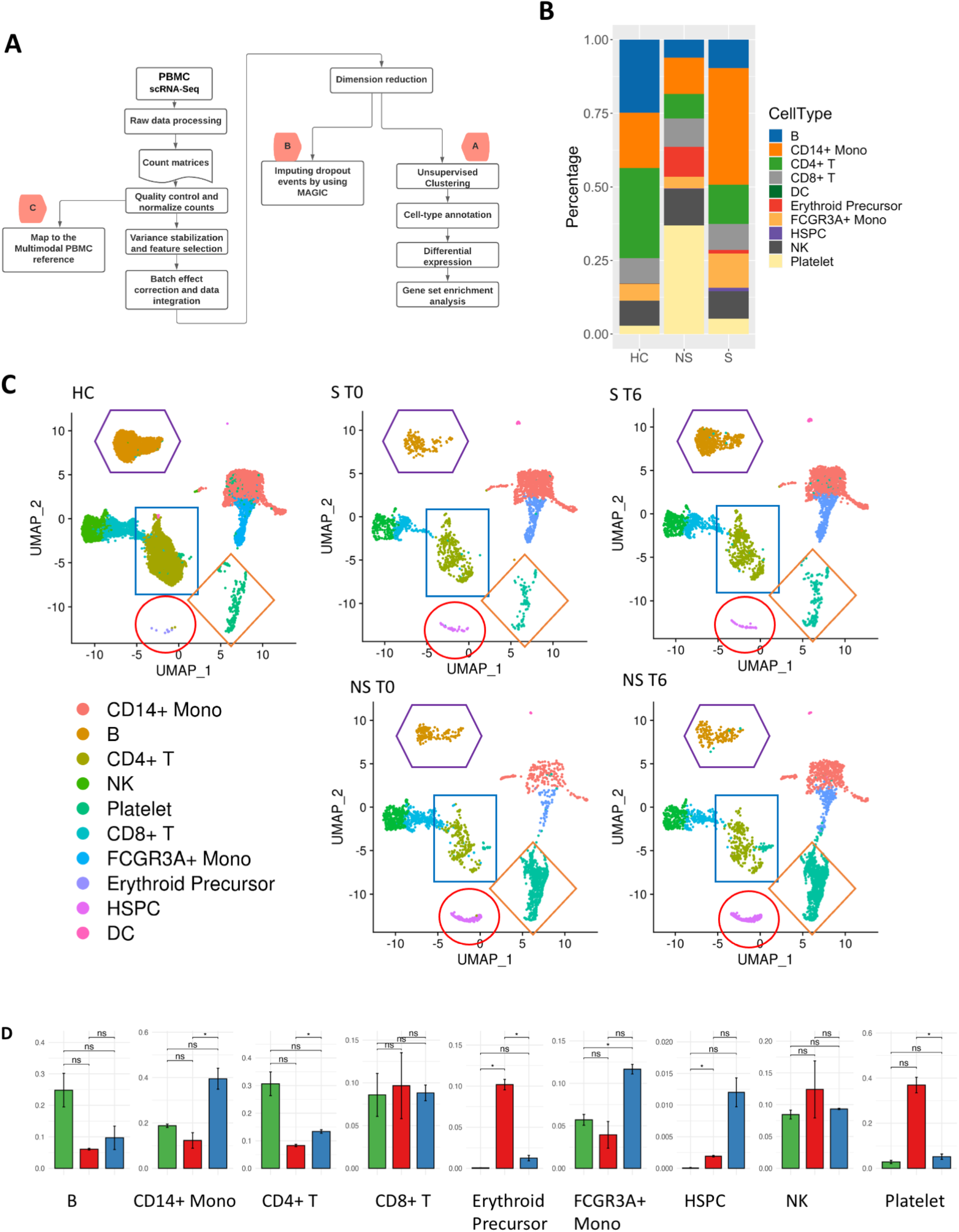
An overview of single-cell transcriptional profiling of PBMC from healthy controls, (HC), survivor (S) and non-survivor (NS) of gram-negative sepsis. **A)** The bioinformatics workflow of the single-cell profiling of PBMC (see Methods section for the detailed description of methods A,B, and C). **B)** Fractions of cell-types in each condition. **C)** The UMAP projection of PBMC from HC, S and NS at 0 hour (T0) and 6 hour (T6) post sepsis recognition. The points representing 27,685 cells are colored according to their types assigned using consensus of three methods (see Methods section). The clusters of cells with the largest changes in sepsis are enclosed in colored shapes: erythroid precursors (red circle), platelets (orange diamond), B cells (purple hexagon) and CD4+ T cells (blue rectangle). **D)** Comparison of cell-type fractions between different conditions (green, red and blue indicate HC, NS and S samples, respectively). Each bar shows an average percentage of a given cell-type across all samples from that condition (only cell types with fractions over 1% across all samples in a condition are shown). Error bars represent ± SEM for 2 HCs and 2 samples from each patient (T0 and T6). HC (n = 2), NS (n = 2) and S (n = 2). The significance was evaluated using two-sample t-tests. The differences with p-values below 0.05 are indicated as *, “ns” – not significant.

The dominant cell types in the healthy controls (HCs) were CD4 T cells (30.45%), followed by B cells (25.04%), and then CD14 monocytes (18.80%). In sepsis survivor (S) and non-survivor (NS), we observed an approximately two-fold decrease in the percentage of CD4 T cells (S, 13.21%; NS, 8.23%) and B cells (S, 10.55%; NS, 6.06%) as compared to the HCs. CD14+ monocytes showed the opposite trend with the highest percentage in sepsis survivor (38.44%) as compared to the HC (18.80%) and the non-survivor (12.82%). These population distributions exhibited similar trends to the flow cytometry analysis (Figure 1), however, we also identified the expansion of atypical PBMC subsets in sepsis. Specifically, we observed a dramatic increase in erythroid precursors and platelets in S and NS PBMC. Only traces of these cell types are typically present following PBMC isolation by gradient centrifugation, therefore their high levels here may suggest abnormal expansion and activation in sepsis. Specifically, HCs contained 2.88% of platelets and 0.07% of erythroid precursors but both subsets were elevated in sepsis, especially in the NS samples (37.43% platelets, 10.28% erythroid precursors) and, to a much lower extent, in the S samples (5.45% platelets, 1.18% erythroid precursors). In general, the populations of most cell types in the non-survivor were more distant from the healthy controls than the cell populations in the survivor. The results obtained with Seurat v3 canonical pipeline were consistent with additional calculations performed with a slightly different methodology using the MAGIC imputation algorithm ^11^ and Seurat v4 map to reference method ^12^. The final assignments of cell types were based on the consensus approach (see Materials and Methods and Supplementary Methods).

We next identified differentially expressed genes (DEGs), and associated pathways (whose down- and upregulations were evaluated using modules’ scores) by comparisons of sepsis samples and healthy controls (Sepsis vs. HCs) and sepsis non-survivor versus sepsis survivor samples (NS vs. S) (Table 2). We also performed temporal comparisons to find DEGs between hour 6 (T6) and hour 0 (T0) (with time counted from sepsis recognition). This temporal analysis was done separately for sepsis non-survivor (NS T6 vs. T0), and sepsis survivor (S T6 vs. T0). Unsurprisingly, the greatest changes were observed between HCs and sepsis patients. CD14+ and FCGR3A+ monocytes exhibited the highest numbers of DEGs (with comparable numbers of down- and upregulated genes) and functional pathway changes in all temporal comparisons. Platelets exhibited the second highest number of DEGs in temporal comparisons, likely related to the striking expansion of this subset in sepsis, especially in non-survivors. In platelets, almost two times more genes were downregulated in sepsis compared to controls than upregulated, suggesting transcription shutdown in sepsis. This trend was also observed when comparing non-survivor to survivor, suggesting that aberrant gene expression and likely transcription shutdown in platelets was indicative of sepsis severity. When comparing gene expression changes within 6 hours within the same patient, there were less DEGs. However, we observed a stark contrast in DEGs for lymphocyte subsets (B cells, T cells, and NK cells); NS exhibited low DEG numbers while S had almost 10-fold more DEGs. This suggests that lymphocyte subsets undergo more transcriptional changes in surviving outcomes, while in fatal sepsis, these cells may be exhausted. The opposite trend was observed in platelets, with more dynamic changes (up and down) occurring in the non-survivor after 6 hours, while there were almost no changes in platelet gene expression in survivors. Taken together, analysis of major changes in gene expression with regards to immune subsets identifies that sepsis leads to a loss in lymphocyte subsets and the expansion of platelets and erythroid precursors, which is more dramatic in fatal sepsis. Additionally, investigation of gene expression over time within the same sepsis patient suggests that fatal sepsis is associated with transcriptional shutdown especially in T lymphocytes, while transcriptional recovery of these immune subsets is observed in surviving sepsis.

**Table 2.** Numbers of differentially expressed genes (DEGs) and enriched or depleted pathways in all analyzed expression comparisons.

| DEGs (FDR < 0.05) |  |  |  |  |
| --- | --- | --- | --- | --- |
| Cell Type | Sepsis vs. HC<br>(Up/Down<br>regulated) | NS vs. S (Up/Down<br>regulated) | NS T6 vs. T0<br>(Up/Down<br>regulated) | S T6 vs. T0<br>(Up/Down<br>regulated) |
| B | 274(142/132) | 357(153/204) | 15(5/10) | 101(43/58) |
| CD14+ Mono | 935(496/439) | 582(335/247) | 298(149/149) | 200(91/109) |
| CD4+ T | 383(182/201) | 386(171/215) | 16(0/16) | 208(92/116) |
| CD8+ T | 686(414/272) | 577(266/311) | 5(0/5) | 78(63/15) |
| Erythroid<br>Precursor | 501(33/468) | 23(11/12) | 1(1/0) | #N/A |
| FCGR3A+<br>Mono | 938(551/387) | 698(343/355) | 110(56/54) | 139(80/59) |
| NK | 587(269/318) | 611(255/356) | 13(4/9) | 187(132/55) |
| Platelet | 1907(690/1217) | 1621(337/1284) | 249(136/113) | 4(1/3) |
| GO-BP & KEGG pathways (FDR < 0.05) |  |  |  |  |
| B | 364(229/135) | 282(92/190) | 256(209/47) | 85(65/20) |
| CD14+ Mono | 1354(718/636) | 565(338/227) | 652(259/393) | 584(220/364) |
| CD4+ T | 340(152/188) | 505(190/315) | 95(0/95) | 260(215/45) |
| CD8+ T | 519(293/226) | 422(104/318) | 17(0/17) | 113(113/0) |
| Erythroid<br>Precursor | 445(66/379) | 153(78/75) | 73(73/0) | #N/A |
| FCGR3A+<br>Mono | 1123(753/370) | 907(475/432) | 361(120/241) | 416(357/59) |
| NK | 668(275/393) | 674(359/315) | 35(4/31) | 226(181/45) |
| Platelet | 1516(585/931) | 1488(331/1157) | 114(63/51) | 168(7/161) |

### MHC class II-related and translation initiation-related genes exhibit distinct sepsis-specific expression patterns

Studies previously reported the decrease of expression of MHC class II-related genes in immune cells in sepsis ^13–16^. We further explored these genes’ expression changes in sepsis, with specific focus on individual cell types, temporal changes within subjects, and association with survival outcomes. We confirmed that MHC class II-related genes including CD74, HLA-DMA, HLA-DMB, HLA-DOA, HLA-DOB, HLA-DPA1, HLA-DPB1, HLA-DQA1, HLA-DQA2, HLA-DQB1, HLA-DQB2, HLA-DRA, HLA-DRB1, and HLA-DRB5 were down-regulated in sepsis as compared to the HCs, especially in the NS samples. While MHC class II-related gene expression pattern in both S and NS samples appear to move closer to the HCs in the period from T0 to T6, the expression of this gene set in S T0 and T6 samples is overall closer to HC’s gene expression (Figure 3A) than the NS samples. Additionally, the temporal trend towards healthy control levels was more apparent in S. For the analysis of differences in MHC class II gene expression in specific cell types, we used the overall MHC Class II module score (Supplementary Figure 2A), which provided a more robust measure than individual DEGs and, thus, more suitable for the smaller number of datapoints. Across most cell types, the MHC class II module score was decreased in sepsis in comparison with HCs, especially in the non-survivor.

**Figure 3.**
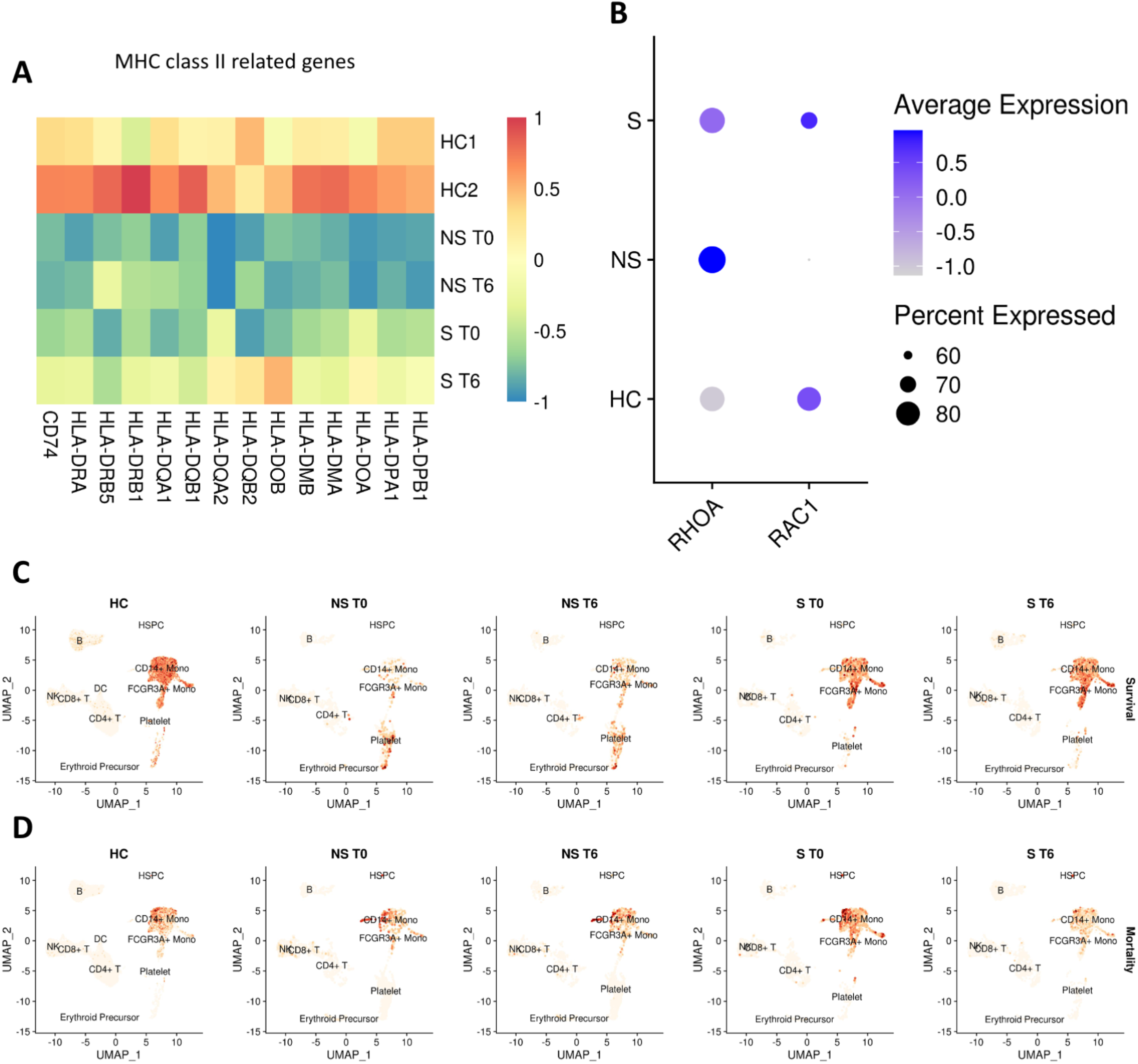
Gene expression modules and survival and mortality signatures across three analyzed conditions (healthy, sepsis survivor and non-sepsis survivor). **A)** The comparison of expression of MHC Class II - related genes in the analyzed samples. Heatmap coloring represents z-scored, log-normalized mean gene expression counts averaged across all cells from a given sample. **B)** The percent of cells with non-zero expression of RHOA and RAC1 genes and average expression of these genes in platelets across three conditions. The color saturation indicates the average expression level, and the circle’s size indicates the percentage of cells expressing a given gene. **C), D)** Expression of genes from **C)** survival and **D)** mortality signatures proposed by Sweeney et.al. ^7^shown in UMAP-based projections of cells distribution in all analyzed samples. Coloring intensity indicates expression levels of each signature.

Average expression of genes related to translation initiation (i.e., genes encoding ribosomal proteins) exhibited a distinctive pattern in sepsis and control samples. In the sepsis survivor (S), these genes, including those encoding L ribosomal, S ribosomal, and mitochondrial ribosomal proteins ^17^, were expressed at a higher level, than in healthy controls and their expression also increased with time (Supplementary Figure 3). The NS samples had the lowest expression score of the module representing these genes across almost all cell types and, consequently, the deviation of this module score from healthy controls was also most noticeable in NS samples (Bonferroni adjusted p-values below 0.01) (Supplementary Figure 2B). The biggest decreases in this module in NS were observed in platelets, CD14+ monocytes, and FCGR3A+ monocytes. These cell types had over 60 ribosomal proteins encoding genes downregulated in NS as compared to S; other cell types had fewer than 15 significantly downregulated genes in the same comparison, ribosome proteins are listed in Supplementary Table 1. Interestingly in monocytes, B cells and CD4+ T cells, the ribosomal genes module score was higher in S samples than in HC, suggesting a mechanism for re-establishment of immune homeostasis in recovery from sepsis. In contrast, the score was lower in platelets, CD8+ T cells, NK cells, and erythroid precursors. These results are consistent with previous studies which have demonstrated that sepsis halts protein translation and hypothesized that this would potentially lead to organ failure ^18, 19^. Our analysis additionally highlights platelets and monocytes as the main cell subsets affected in fatal sepsis outcomes, where they exhibit translation shutdown within hours.

### Aberrant platelet gene expression is observed in fatal sepsis

The role of platelets in the development of sepsis pathophysiology is not well established. Still, increasing number of studies are beginning to recognize that platelets are altered in sepsis, and that transcriptional and translational changes in platelets are related to mortality ^20^. Out of all the cell types analyzed, platelets contained the highest number of DEGs in our comparisons of sepsis samples vs. HCs and in comparisons of NS vs. S samples. Specifically, the number of DEGs in platelets from both comparisons was more than two-fold higher than in FCGR3A+ monocytes, which ranked second in the number of DEGs in these comparisons (Table 2). While most DEGs in platelets were down-regulated in sepsis, genes contributing to microvascular coagulation were up-regulated (64% in Sepsis vs. HC, and 79% in NS vs. S, see Supplementary Table 1 for the list of genes included in coagulation module). Sepsis non-survivor samples had higher coagulation module scores than HC and S samples in most cell types, but this trend was most pronounced in platelets (Supplementary Figure 2C). In contrast, in survivor samples, only platelets exhibited higher coagulation module scores than HC.

In platelets, significant gene expression changes were observed for the Rho GTPases RAC1 and RHOA, which regulate cell adhesion ^21^. RAC1, the main GTPase required for cell barrier maintenance and stabilization was downregulated in NS vs. S comparison (logFC of 0.31 and adjusted p-value < 0.001). RHOA, the GTPase that negatively regulates barrier properties under both resting and inflammatory conditions was up-regulated in Sepsis vs. HC (logFC of 0.6 and adjusted p-value < 0.001) (Figure 3B). In NS, the expression changes in platelets suggest both increased microvascular permeability and microvascular coagulation. These phenomena may contribute to the rapid development of multi-organ failure.

The genes overexpressed in platelets in Sepsis vs. HC and NS vs. S comparisons included MHC class I-related genes such as HLA-A, HLA-B, HLA-C, HLA-E, and HLA-F (Supplementary Figure 2D). In Sepsis vs. HC comparisons these genes have logFC values between 0.3 and 1.1, while in NS vs. S comparisons the logFC values were 0.3-0.8, all with adjusted p-values < 0.001. In the NS the score for MHC Class I expression module also reached the highest values (Supplementary Figure 2D). Platelets are reported to express MHC class I, which is significantly increased during infection both on their plasma membrane and intracellularly ^22, 23^, a phenomenon that may affect CD8+ T cell responses by platelet MHC class I antigen cross-presentation.

The expression modules related to responses to type I IFN, IFN-gamma, and IFN-beta also had the highest scores in platelets in comparison of NS with other samples (Supplementary Figure 2E-G). Overall, these dramatic changes in platelet gene expression related to coagulation, CD8 T cell modulation and inflammation suggest that aberrant platelet function may be predictive of sepsis disease severity and fatality through effects on the vasculature and immune cells.

### Monocyte gene changes are associated with sepsis disease severity

The published studies reporting biomarker genes in sepsis were mostly based on bulk data from all immune cells from the peripheral blood. The extensive study by Sweeney et al. ^7^ used a community-based approach to establish optimized lists of genes whose expressions have the strongest prognostic value for sepsis mortality and survival. We tested if these lists could be used in single-cell transcriptomic data to provide insights into cell types crucial for sepsis outcomes. Interestingly, genes linked to sepsis survival were mostly expressed in monocytes from HC and S T6 samples (Figure 3C). On the other hand, the genes linked to sepsis mortality were more represented in monocytes from NS and S T0 samples (Figure 3D).

Next, we evaluated monocyte-specific cytokine expression. Compared to HCs, CD14+ monocytes from sepsis patients were characterized by upregulation of chemokines (CCL3 and CCL4) (Figure 4A). Pro-inflammatory cytokines, chemokines and adipokines including IL6, CCL2, CCL3, CCL4, CCL7, HMGB1, and NAMPT were overexpressed in NS as compared to S samples (Figure 4B). Amphiregulin (AREG) was also overexpressed in NS, where it may have a pro-inflammatory role through promoting production of pro-inflammatory cytokine IL8 (CXCL8) ^24^. CXCL8 itself was upregulated in NS as compared to S in FCGR3A+ monocytes (Figure 4D). Another up-regulated gene - EBI3, is also reported to promote pro-inflammatory IL6 functions by mediating trans-signaling ^25^. Of note, PPBP (CXCL7), a potent neutrophil chemoattractant also expressed by platelets ^26^, was one of the top genes upregulated in both monocyte subsets and in both comparisons (Sepsis vs HC, and NS vs S). These results suggest that monocytes in sepsis are in hyperinflammatory state, which is more severe in patients with fatal outcomes.

**Figure 4.**
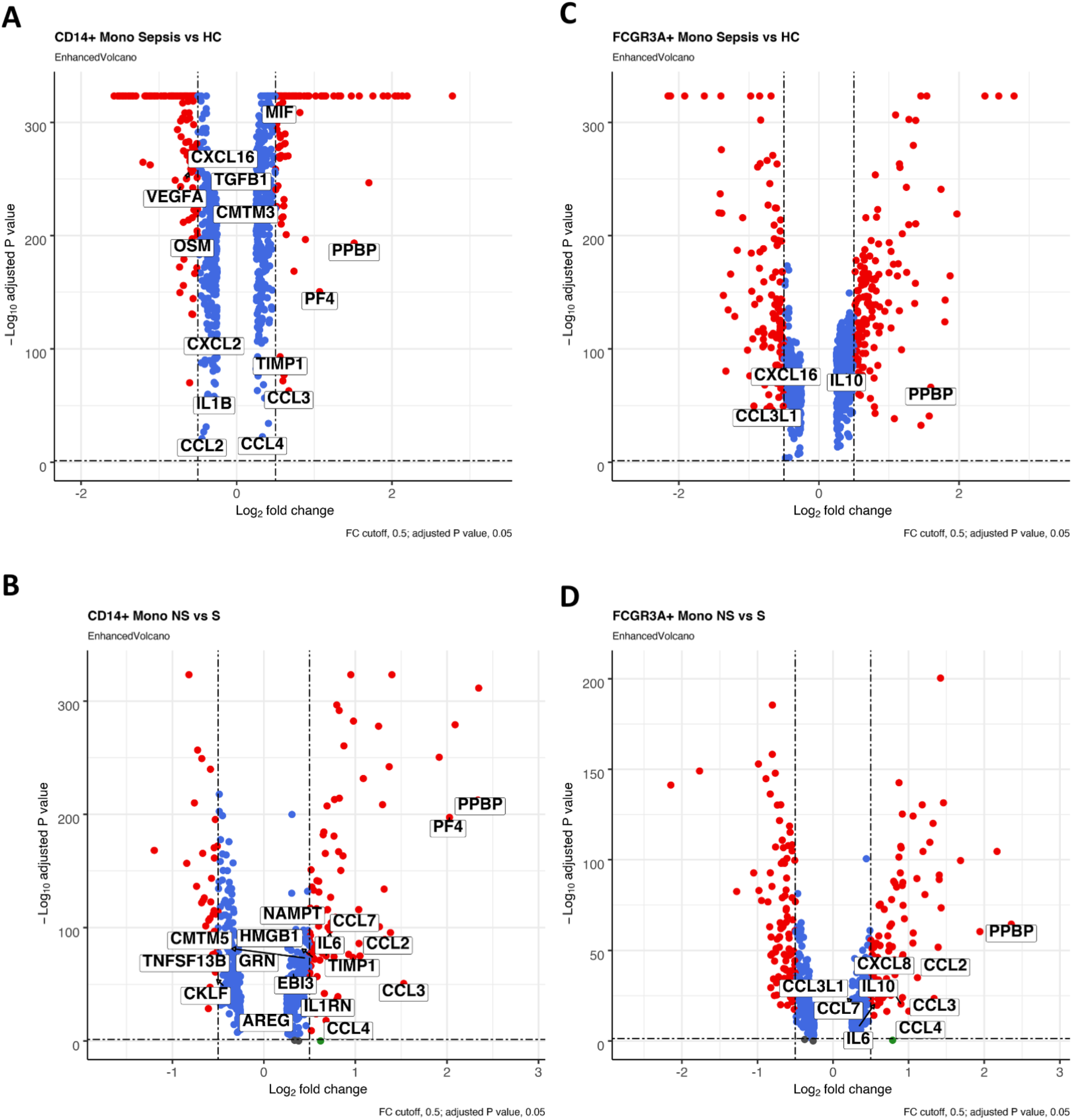
Expression changes of genes encoding cytokines in specific cell-types illustrated by volcano plots. **A)** Expression in CD14+ monocytes from sepsis versus HC; **B)** CD14+ monocytes from NS vs. S; **C)** FCGR3A+ monocytes from sepsis vs. HC; **D)** FCGR3A+ monocytes from NS vs. S; Each dot represents one gene, red color indicates significantly overexpressed genes (LogFC > 0.5, and FDR adjusted p-value < 0.05); blue indicates significantly under expressed genes (0 < LogFC < 0.5, and FDR adjusted p-value < 0.05); green indicates genes with LogFC > 0.5, but FDR adjusted p-value > 0.05; gray indicates genes without significant expression differences. Cytokines (i.e. genes included in Gene Ontology term “cytokine activity” - GO:0005125) are labeled. Volcano plots were prepared with R package EnhancedVolcano ^54^.

### Sepsis survival is associated with increased IFN response and reduced exhaustion in cytotoxic lymphocytes

The comparison of CD8+ T and NK cells in samples across the three conditions showed that, in cytotoxic cells, the sepsis survivor (S) had the highest IFN response score, including type I IFN, IFN-γ, IFN-β responses (Supplementary Figures 2E-G). Conversely, in the same cell types, the sepsis non-survivor (NS) had the lowest module score of response to IFN-gamma (Supplementary Figure 2F), suggesting that hyporesponsiveness and lymphocyte exhaustion is associated with fatal outcomes. Independently from that, we assessed lymphocyte exhaustion level using the exhaustion module comprised of co-inhibitory receptors such as PDCD1, HAVCR2, LAG3, CD244, ENTPD1, CD38, CD101, TIGIT, and CTLA4 ^27^, and regulators downstream of TCR signaling pathway such as TOX, NR4A1, and IRF4 ^28^. The results indicated exhaustion of NK and CD8+ T cells in sepsis patients as compared to HC (Supplementary Figure 4A). Notably, in NK cells the NS samples had a much higher exhaustion score than S samples (Supplementary Figure 4B).

We investigated T cell-mediated cytotoxicity using a list of genes associated with GO term “T cell mediated cytotoxicity” (GO:0001913). Compared to the HCs, sepsis patients had higher expression of genes associated with T-cell cytotoxicity in CD8+ T cells (Supplementary Figure 4C). However, when comparing NS and S samples, we observed that CD8+ T and NK cells from NS had significantly lower expression of cytotoxicity-related genes than S (Supplementary Figure 4D), confirming the exhaustion and immunosuppressed state of lymphocytes in NS.

Taken together, these transcriptomic findings indicate that NK and T lymphocytes in the sepsis survivor had a more robust response to IFNs. Although all CD8+ T and NK cells in the sepsis samples exhibited gene expression signatures associated with exhaustion, the sepsis survivor had adaptive cytotoxic lymphocytes activated, while the non-survivor exhibited reduced cytotoxic activity in both innate and adaptive lymphocyte subsets. These findings support the predictive potential of lymphocyte exhaustion for detrimental outcomes in sepsis. Consistent with other studies, our findings also support the therapeutic potential of inhibiting lymphocyte apoptosis and restoring lymphocyte function for new treatments to restore immune homeostasis after sepsis episodes ^29–34^.

### Sepsis drives metabolic shift and hypoxic stress which is exacerbated in fatal outcomes

The metabolic state of specific immune cells profoundly determines their function. In the steady-state, immune cells rely on oxidative phosphorylation (OXPHOS) for ATP production as the energy source, maintaining homeostasis. In sepsis, immune cells such as monocytes undergo the process known as Warburg effect, in which the metabolism mechanism shifts from OXPHOS to glycolysis by using lactic acid fermentation as the predominant energy source ^35^. The transcription factor hypoxia-inducible factor (HIF1A) is a main driver for this metabolic switch, of relevance to the hypoxemia in severe sepsis ^36^. We examined HIF1A expression among cell types across the three conditions (HC, NS, and S). CD14+ monocytes expressed the highest HIF1A levels, followed by FCGR3A+ monocytes. Most monocytes from the sepsis non-survivor expressed higher levels of HIF1A than monocytes in the sepsis survivor, where expression of HIF1A was comparable to healthy controls (Supplementary Figure 5A).

**Figure 5.**
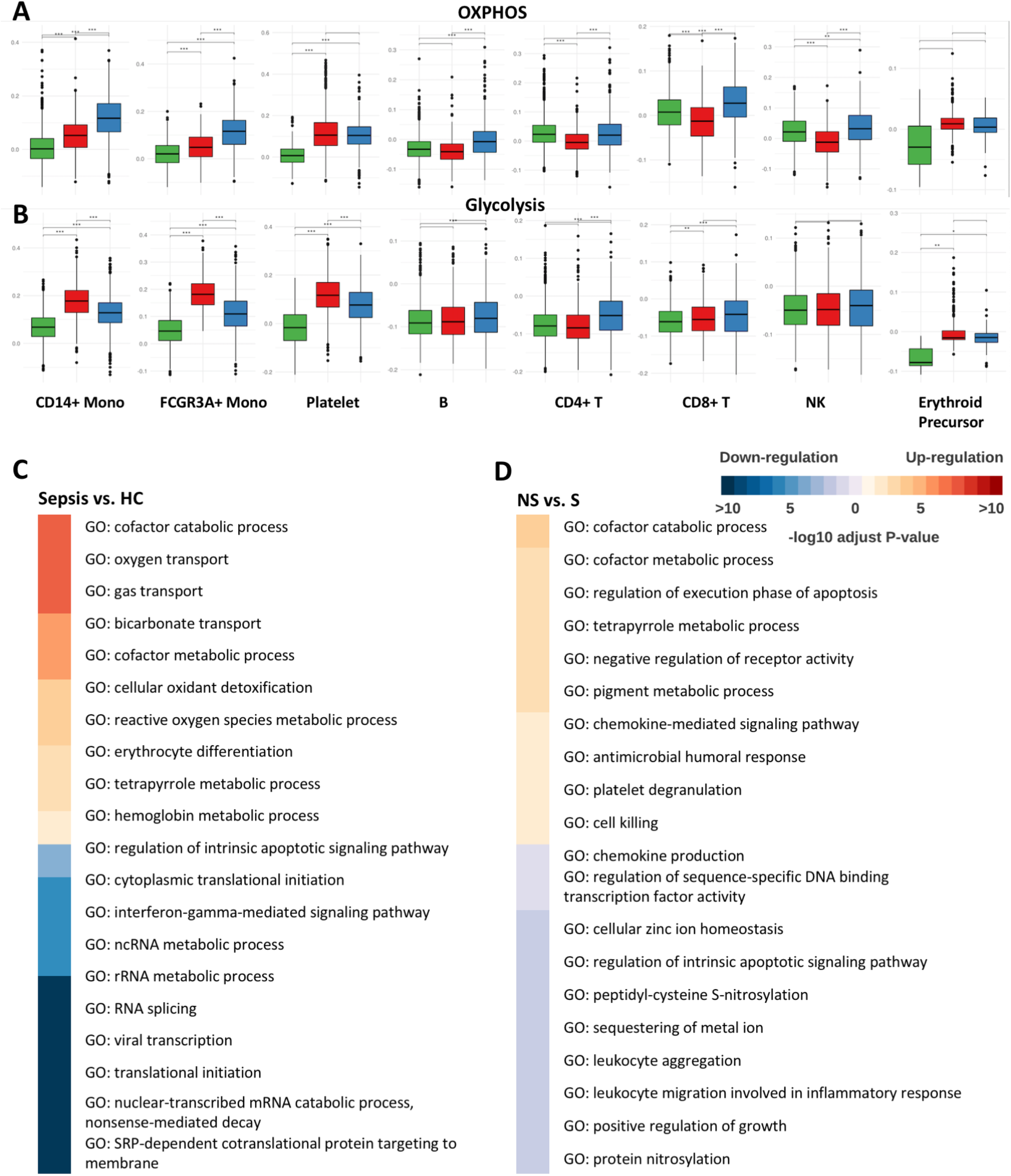
Comparisons of immune cells from healthy controls versus sepsis non-survivor and survivor. **A)** Oxidative phosphorylation (OXPHOS) module and glycolysis module **B)** scores of different cell types from each sample. The differences with Bonferroni adjusted p-values below 0.01, 0.001, and 0.0001 are indicated as *, **, and ***, respectively. The significance analysis was performed using two-sample t-tests. **C), D)** Pathway enrichment when comparing erythroid precursors **C)** sepsis vs. HC, **D)** NS vs. S. All the GO terms were reduced to representative ones by Revigo ^55^with similarity at 0.4, then the top 10 −log10 adjust P-value were selected shown in the heatmap.

We next evaluated scores of OXPHOS and glycolysis modules in different cell-types using gene lists reported by Frederick et al. ^37^. In CD14+ monocytes, FCGR3A+ monocytes, B cells, CD8+ T cells, and NK cells from sepsis survivor, the scores of OXPHOS module exceeded the corresponding scores in the NS and HC samples. At the same time in CD4+ T cells, CD8+ T cells, and NK cells from sepsis non-survivor, OXPHOS module scores were below the corresponding scores in S and HC samples (Figure 5A, Bonferroni adjusted p-value < 0.001). This reduction in OXPHOS in the lymphocytes of sepsis non-survivors is consistent with the previous observation of lymphocyte exhaustion in fatal sepsis ^38^. At the same time, the glycolysis module score was observed in multiple cell types from the sepsis non-survivor to significantly exceed values in NS and HC samples (CD14+ monocytes, FCGR3A+ monocytes, and platelets, Bonferroni adjusted p-value < 0.0001). None of the cell types from the sepsis survivor appeared in the highest module score in glycolysis across three conditions (Figure 5B). These results indicate that in severe sepsis, monocytes and platelets shift their metabolic function from OXPHOS to glycolysis, and this shift is more extreme in fatal outcomes. Interestingly, in sepsis platelets showed increased module scores for both OXPHOS and glycolysis, suggesting metabolic dysfunction and competition for energy sources. In healthy controls, monocytes and platelets had OXPHOS and glycolysis module scores significantly below levels observed in both sepsis samples (Bonferroni adjusted p-value < 0.001) (Figure 5A, B), confirming that monocytes and platelets have dysregulated metabolic activity in sepsis patients.

To investigate if HIF1A was the main driver of the observed metabolic changes, and to evaluate which cell types were responsive to HIF1A, we evaluated correlations between HIF1A expression and OXPHOS/glycolysis module scores. Cells with no detected HIF1A expression were filtered out, resulting in 14,077 cells suitable for the analysis. Across the three conditions (HC, S and NS), where significant correlation between HIF1A and OXPHOS was observed (correlation p-value < 0.05), the correlation was negative in CD14+ Mono, FCGR3A+ Monocytes, and platelets, which had the absolute correlation coefficient |R| above 0.25 and p-value < 0.05 (Supplementary Figure 5B). The similar analysis of correlation between HIF1A expression and glycolysis showed the opposite effect where CD14+ Mono, FCGR3A+ Monocytes, and platelets across the three conditions with significant correlation (p-value < 0.05) had a positive correlation coefficient. The highest correlation coefficients, exceeding 0.25, were observed CD14+ and FCGR3A+ monocytes from the sepsis non-survivor (Supplementary Figure 5C).

The erythroid precursor expansion observed in the non-survivor sepsis patient was also indicative of severe hypoxemia in fatal sepsis ^39^. We compared pathway changes in sepsis vs. HC and NS vs. S. Erythroid precursors in sepsis expressed genes related to hypoxic stress (oxygen transport, erythrocyte differentiation, response to oxidative stress, cofactor catabolic process and cellular oxidant detoxification) (Figure 5C). The pathways that were down-regulated in sepsis vs. HC and NS vs. S (Figure 5C–D), included translational initiation, RNA splicing, protein nitrosylation and regulation of sequence-specific DNA binding transcription factor activity, suggesting that erythroid precursors in sepsis exhibited a halt in protein translation.

Together, these data suggest that the peripheral immune cells in sepsis are responsive to the hypoxic environment that leads to HIF1A-mediated shifts in metabolic function towards glycolysis, with fatal outcomes exhibiting more extreme hypoxic stress leading to protein shutdown especially in erythrocyte precursors. These results support the possibility of therapeutically targeting HIF1A to restore metabolic homeostasis in the sepsis immune response.

### Temporal changes in the transcriptome profile elucidate differences between the survivor and the non-survivor in the first hours of sepsis recognition

To reveal trends associated with recovery and fatal outcomes we analyzed temporal changes of gene expression in immune cell subsets during the first 6 hours from sepsis recognition. When comparing hour 6 and hour 0 in sepsis non-survivor (NS T6 vs. T0), the cell type that had the most differentially expressed genes (DEGs) and pathway changes were the CD14+ monocytes. For the sepsis survivor (S T6 vs. T0 comparison), the cell type with the highest number of DEGs were the CD4+ T cells, and the cell type with the highest number of pathway changes were the CD14+ monocytes (Table 2). To examine how CD14+ monocytes contribute to the fatal sepsis outcome, we investigated functional pathways for which expression was increased in non-survivor (NS T6 vs. T0) but decreased in survivor (S T6 vs. T0) (Figure 6A). One of the GO pathways associated with fatal outcomes appeared unrelated to sepsis (‘female pregnancy’). However, the genes in this pathway were involved in tissue remodeling and fibrosis (eg. CALR/TIMP1/ADM), which are also related to the “inflammatory response to wounding pathway) potentially suggesting vessel disruption and remodeling in fatal sepsis. Additionally, pathways related to metabolic dysfunction, and inflammatory and oxidative stress were also increased over time in fatal sepsis. To examine the role of CD14+ monocytes in sepsis recovery we tested which pathways’ expression was decreasing in sepsis non-survivor (NS T6 vs. T0) but increasing in sepsis survivor (S T6 vs. T0). Those functional pathways, likely associated with recovery, included cell migration, and regulation of inflammatory response (Figure 6E). The above results suggest that within 6 hours of sepsis progression the CD14+ monocytes in the non-survivor undergo increasing response to cellular stress and inflammation and trigger tissue remodeling processes, while exhibiting waning functions of neutrophils, cell migration, and reduced immune cell proliferation. FCGR3A+ monocytes also exhibited multiple pathways with opposite temporal expression trends in non-survivor and survivor (NS T6 vs. T0 and S T6 vs. T0, Figure 6B). The only significant pathway which increased over time in fatal sepsis was regulation of chemokine (genes up in NS T6 are PYCARD/GSTP1/CSF1R, and genes down in S T6 are HIF1A/CLEC7A/DDX3X), which may suggest that this regulatory monocyte subset in non-survivors was more immunosuppressed and/or refractory to outside chemotactic signals. In the FCGR3A+ monocytes from the sepsis survivor, there was an increase of genes over time related to monocyte migration, cell-cell interaction and metabolic activity, as well as regulation of apoptosis. These suggest a return to homeostasis and normal monocyte function and increased cell survival.

**Figure 6.**
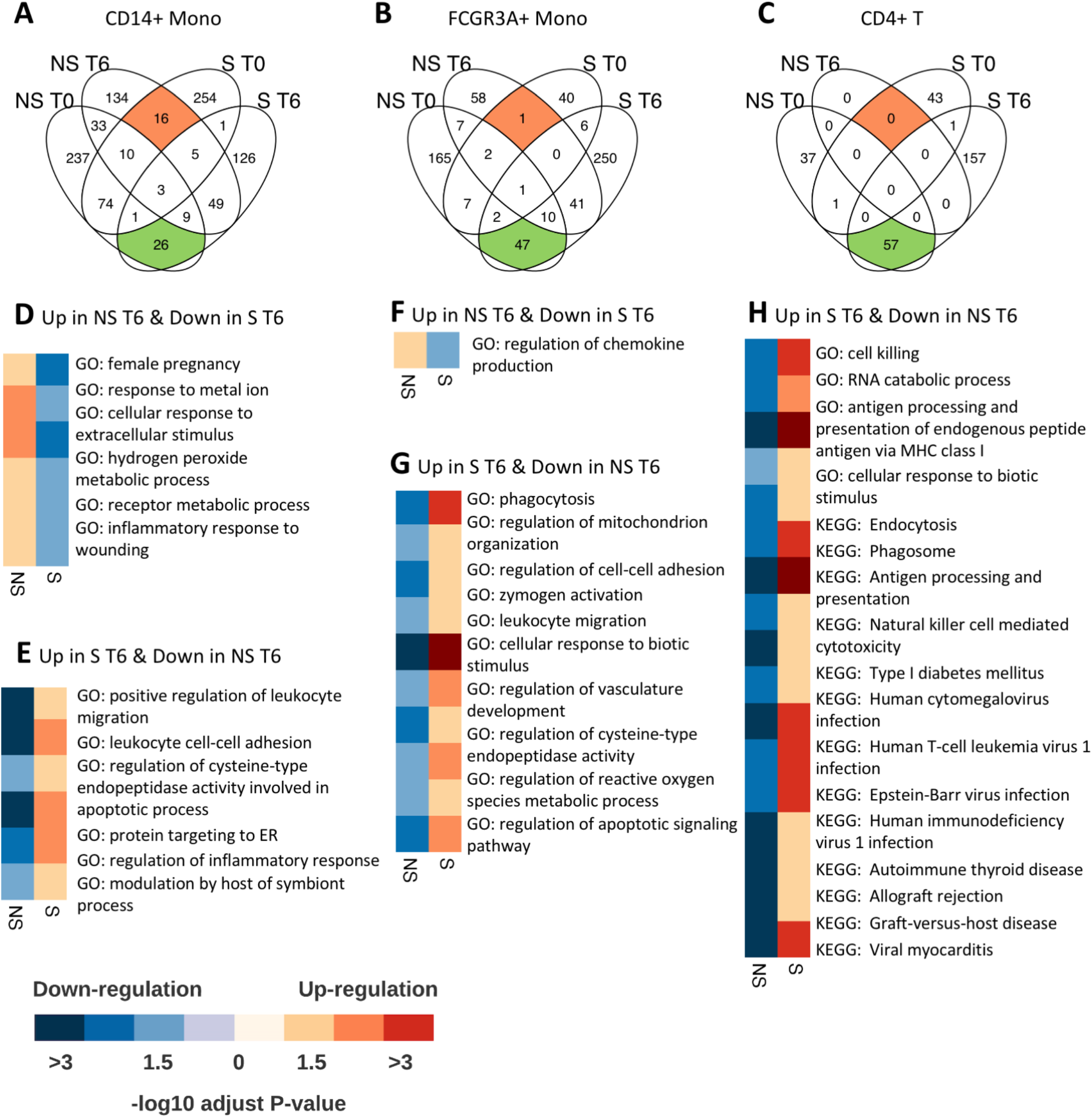
Opposite temporal changes in pathway expression between sepsis non-survivor and survivor. Venn diagrams describing temporal changes in pathways from each sepsis patient. Set labeled ‘NS T0’ contains pathways decreasing in NS between T0 and T6, set labeled ‘NS T6’ contains pathways increasing between T0 and T6 in NS, etc. **A)** CD14+ monocytes, **B)** FCGR3A+ monocytes, **C)** CD4+ T cells. Sets of pathways increasing in NS (up-regulated in NS T6 as compared to NS T0) and decreasing in S (down-regulated in S T6 as compared to S T0) is colored in orange. Set of pathways decreasing from NS T0 to T6 and increasing from S T0 to T6 re colored in green. **D)** Heatmap illustration of temporal changes of pathways which are changing in opposite direction in NS and in S **D),E)** in CD14+ monocytes, **F),G)** in FCGR3A+ monocytes, **H)** in CD4+ T cells. The sets of overlapping GO terms were reduced to representative ones using Revigo ^55^ (the cutoffs were, more than 10 overlapping GO terms and similarity > 0.4).

Among lymphocytes, CD4+ T was the cell type that had the highest number of pathways showing opposite temporal trends in survivor and non-survivor (NS T6 vs. T0 and S T6 vs. T0, Figure 6C). Interestingly, all these pathways were decreasing in the CD4+ T cells from non-survivor and increasing in the survivor (Figure 6G). The functions of these pathways, possibly associated with recovery, are related to T cell cytotoxic function including and response to pathogens. This suggests that the rapid recovery of T cells (in particular, their reversal from exhaustion) may be crucial for positive outcomes in sepsis.

Other cell types where pathways showed opposite temporal trends in non-survivor and survivor (NS T6 vs. T0 and S T6 vs. T0) were the CD8+ T cells, NK cells, and platelets (Supplementary Figures 6A-C). CD8+ T cells and NK exhibited trends similar to CD4+ T cells, where all pathways with opposite trends were decreasing in non-survivor (NS T6 vs. T0) but increasing in survivor (S T6 vs. T0). The functions of these pathways were mainly related to protein synthesis (Supplementary Figures 6D, E). In platelets, the pathways increasing non-survivor (NS T6 vs. T0) but decreasing in survivor (S T6 vs. T0) involved apoptotic, metabolic, and protein folding processes (Supplementary Figure 6F).

In summary, the analysis of single-cell transcriptomics in sepsis with different outcomes revealed distinct dynamic trends in expression in immune cell subsets, highlighting the benefits of tracking temporal changes by single cell analysis, which could be specifically targeted to improve outcomes. In monocytes, the fatal outcome appears to be associated with increasing expression of pro-inflammatory pathways but, at the same time, with the inability to respond to external stimuli. Similarly, in CD4+T cells, increasing exhaustion and hyporesponsiveness were associated a fatal outcome. For the other PBMC subsets, pathways correlated with a fatal outcome were not immune-specific, but rather associated with protein production shutdown, that is reflective of dysfunction and exhaustion, as observed in the monocytes and T cells, respectively.

## Discussion

Sepsis is a dysregulated systemic inflammatory response, which alters the innate and adaptive immune responses to microbial invasion and results in organ injury with mortality rates of 15–25% ^40, 41^. Given significant sepsis disease heterogeneity, single cell transcriptomics offers an valuable approach to provide a better understanding of the molecular mechanisms of sepsis, however, from over 1000 single-cell transcriptomics studies have been published to date^42^, only two studied sepsis ^8, 43^. In those two studies, the authors focused on monocytes and myeloid-derived suppressor cells by sorting on specific surface markers. In comparison, our study used the centrifuge gradient-based approach before performing the single-cell RNA-seq, which expanded the cell subsets investigated in the study. We additionally collected samples at different time points from a survivor and non-survivor of sepsis, which provided temporal details of the immune response in severe sepsis. Limitations of this study include the small sample size, therefore, we focused on two dramatically different clinical outcomes (fatal versus recovery), while using a within-subject study design where patients had similar clinical syndrome and sepsis etiology. In these focused analyses were able to identify specific immune cell subsets and gene expression patterns over time that correlated with beneficial or fatal outcomes.

Our results are consistent with the previous studies both in single-cell and bulk sepsis transcriptomic studies. The peripheral blood cell composition of non-survivors is much more “distant” from healthy controls than the blood cells of survivors (Figure 2B–D). In addition, we identify cell types that were not studied by the previous single-cell studies, which include platelets and erythroid precursors. These were found to be expanded in sepsis patients, especially in the sepsis non-survivor. Examination of platelets from the non-survivor sepsis patient revealed increased expression of genes related to microvascular permeability and microvascular coagulation, which in the literature were reported to lead to the development of organ failure ^21^. The erythroid precursors, that were dramatically expanded in non-survivors, showed upregulation of genes related to hypoxic stress and apoptosis, reflective of the hypoxic environment in severe sepsis that leads to emergency erythropoiesis. Interestingly, according to a longitudinal COVID-19 study ^44^, erythroid cells were identified as the hallmark of severe disease and had defined molecular signatures linked to a fatal COVID-19 disease outcome. Here we also observed that erythrocyte expansion and expression of genes related to hypoxic stress was a major predictor of fatal outcomes.

Consistent with previous studies showing that CD14+ monocytes play a significant role in the sepsis pathogenesis ^45, 46^, we observed aberrant gene expression and pathway changes in the monocytes of sepsis patients, suggesting they were in a hyperinflammatory state (Table 2, Figure 3C–D). However, in a novel observation, monocytes from sepsis patients, especially the non-survivor, exhibited signs of being refractory to the external environment, with reduced expression of interferon responsive genes and the inability to produce TNFα in response to LPS treatment (Table 1) (Supplementary Figure 2E, F, G). The lymphocytes in the sepsis patients also showed reduced lymphocyte-mediated functions, with the sepsis survivor showing less lymphocyte exhaustion performance but maintaining their cytotoxic function (Supplementary Figure 4C-D). When we focused on the metabolic pathway changes, we found that monocytes and platelets from the sepsis non-survivor exhibited a switch in their energy utilization, from the OXPHOS to partial OXPHOS and glycolysis (Figure 5A–B). At the same time, the HIF1A expression and metabolic function pathway correlation analyses suggested that HIF1A was the main driver of changing the metabolic pathways (Supplementary Figure 5B-C).

By exploring the temporal changes in the transcriptome profile from the sepsis non-survivor and the survivor, we found dramatic differences predominantly involving CD14+ monocytes (Figure 6A). 6 hours following sepsis recognition, the non-survivor’s CD14+ monocytes demonstrated an intense response to stimulation, stress, and inflammatory behaviors. In contrast, these pathways were downregulated in the sepsis survivor during this time (Figure 6D). Further, the survivor’s CD14+ monocytes expressed pathways related to cell migration, neutrophil function, lymphocyte proliferation, while these were downregulated in the non-survivor (Figure 6E). Overall, our study suggests that not only the initial status of the sepsis patient but also the dynamic changes in cell behavior during the critical period following diagnosis significantly effect sepsis outcome. Future focus on these changes, specifically addressing immune cell metabolic dysfunction and identifying mechanisms to promote their recovery from exhaustion may provide therapeutic and prognostic insight into sepsis.

## Materials and methods

### Human blood collection and harvest of PBMCs

#### Enrollment

Human peripheral blood was collected from non-sepsis donors from the Riverside Free Clinic and septic patients with signed informed consent and approval of the University of California, Riverside (UCR, #HS-17-707), and Riverside University Health System (RUHS, #1024190-3) Institutional Review Board. Sepsis patient enrollment was performed according to the following inclusion criteria: 1/Admission to Intensive Care Unit; 2/Age greater than or equal to 18 years old; 3/Suspected or confirmed infection; 4/qSOFA score ⩾ 2 (qSOFA variables: altered mentation [GCS ⩽13], systolic blood pressure < 100 mmHg and respiratory rate > 22 breaths/minute) and/or; 5/Lactate greater than or equal to 2.0 mmol/L and on vasopressor therapy to maintain MAP > 65 mmHg after 30 mL/kg intravenous fluid bolus.

#### Peripheral blood mononuclear cells (PBMC) analysis

Blood was recovered in Vacutainer glass collection tubes with heparin (BD Biosciences). PBMC were isolated by gradient centrifugation with Histopaque-1077. Plasma was recovered for cytokine quantification by cytokine bead array (BD Biosciences), and resistin ELISA, (Peprotech). Cell aliquots were frozen in liquid nitrogen. Following blood draw, PBMC isolation was performed within 24 hours through density gradient centrifugation and cells were stored immediately in liquid nitrogen. Flow cytometry characterization of PBMC involved incubation with Human TruStain FcX™(Biolegend), and staining with primary antibodies: CD14(HCD14, Biolegend), CD16(3G8, Biolegend), CD66b(G10F5, eBioscience), CD3(OKT3, eBioScience). Samples were acquired on a BD LSRII and analyzed on FlowJo (v10).

#### 10X genomics

For single cell sequencing, thawed PBMC live cells were recovered by centrifugation-based dead cell removal kit (Miltenyi) and viable cells confirmed by hemocytometer counting (>85% viable). 15,000 cells per sample were loaded onto the 10x genomics platform, and cDNA libraries prepared according to manufacturer’s instructions (Chromium Next GEM Single Cell V3.1). Samples were sequenced at UCSD Genomics center on the NovaSeq platform at 250M reads/sample.

### Process and quality control of the single-cell RNA-seq data

The Cell Ranger Software Suite (v.3.1.0) was used to perform sample de-multiplexing, barcode processing, and single-cell 5’ unique molecular identifier (UMI) counting. Specifically, splicing-aware aligner STAR was used in FASTQs alignment. Cell barcodes were then determined based on the distribution of UMI counts automatically. The following criteria were applied to each cell of four sepsis samples and two healthy controls: gene number between 200 and 6,000, UMI count > 1,000, and mitochondrial gene percentage < 0.2. After filtering, a total of 27,685 cells were left for the following analysis. Finally, all samples’ filtered gene-barcode matrix was integrated with Seurat v.3 ^10^ to remove batch effects across different samples.

### Dimensionality reduction, clustering and consensus-based cell type annotation

The filtered gene barcode matrix was normalized using ‘LogNormalize’ method from Seurat package v.3 with default parameters. In the next step, the vst method implemented in the FindVariableFeatures function of the Seurat package was applied to find the top 2,000 most variable genes. It was followed by the principal component analysis (PCA) and the application of the uniform manifold approximation and projection (UMAP) algorithm for cell data visualization performed based on the top 50 principal components. Then the graph-based clustering was performed by applying the FindClusters function of the Seurat package on the PCA-reduced data. With the resolution set to 1, 27,685 cells were grouped into 21 clusters. The first method of assignment of cell types to cell clusters was based on their canonical markers: B cells (MS4A1 marker), CD14+ monocytes (CD14 and LYZ), CD4+ T cells (IL7R, CCR7, and CD27), CD8+ T cells (CD8A), DCs (FCER1A, CST3, CD123, and GZMB), erythroid precursors (GYPB and AHSP), FCGR3A+ monocytes (FCGR3A and MS4A7), Neutrophils (JAML and SERPINB), NK cells (GNLY and NKG7), and Platelets (PPBP). Independently from this initial marker-based cell type assignment we applied cell-type annotation tools SingleR ^47^ and scCATCH ^48^. The SingleR program first identifies genes with big variation between cell types in the reference data set, then compares each cell’s scRNA-seq data with each sample from the reference data set and, lastly, performs iterative fine-tuning to select the most likely cell type of each cell. The microarray dataset from Human Primary Cell Atlas Data with assigned labels was used as the reference. Finally, each cluster was assigned a cell type with the highest percentage of cells assigned to that type by SingleR. The third applied method of cell type assignment was scCATCH, where cell types are assigned using the tissue-specific cellular taxonomy reference databases ^49–51^ and the evidence-based scoring protocol. Our final assignment of cell types to clusters was based on the consensus of the three aforementioned methods as follows: First, each cluster was assigned a cell type selected by most methods if possible. If each method gave a different result, then the priority was given to the assignment based on canonical markers. If the markers-based assignment was inconclusive, the consensus assignment was based on the results from SingleR method.

Independently from the main computational pipeline described above, we applied two alternative pipelines: 1) Single-cell RNA-seq data imputation with the MAGIC algorithm with the default settings ^11^. 2) The Seurat v.4 package which allows skipping the steps of cell clustering and cell annotation, by mapping scRNA-seq sctransformed normalized data directly to the CITE-seq reference of 162,000 PBMCs (FindTransferAnchors and MapQuery functions of Seurat v.4) ^12^. As described in the Results section, the results from these three pipelines were mostly consistent.

### Differential gene expression analysis and functional annotation of genes

The MAST method ^52^ from the Seurat v.3 package (implemented in FindAllMarkers function) was used with default parameters to perform differential gene expression analysis. Differentially expressed genes (DEGs) were found by performing the following comparisons: sepsis samples to HC (healthy controls), sepsis NS (non-survivor) samples to S (survivor) samples, NS samples from T6 (hour 6) to NS samples from T0 (hour 0 i.e., sepsis recognition), and S samples at T6 to S T0. A difference in gene’s expression was considered significant if an adjusted p-value was below 0.05. The false discovery rate (FDR) adjustment was performed by MAST. Only genes with FDR-adjusted p-values < 0.05 were considered in the second step of DEG analysis where we analyzed differences between the results of the comparisons listed earlier. Pathway enrichment analysis was performed by clusterProfiler ^53^ using database Gene Ontology biological process terms (GO-BP) and Kyoto Encyclopedia of Genes and Genomes (KEGG) pathways. The clusterProfiler program was used for statistical analysis and visualization of functional profiles for DEGs with FDR-adjusted p-value < 0.05. The results of differential gene expression and functional enrichment are summarized in Table 2.

### Comparison of module scores

We used cell’s module scores as a measure of the degree at which individual cells expressed certain predefined expression gene sets. The AddModuleScore function from the Seurat v.3 package with default settings was used to perform all calculations and comparisons of module scores. We compared expression of modules such as MHC class II, Ribosomal proteins, Coagulation, MHC class I, Response to type I IFN, Response to interferon-gamma, Response to interferon-beta, Exhaustion, Cytotoxicity, OXPHOS, and Glycolysis. The lists of genes defining these modules were prepared based on Gene Ontology and literature. Genes without detectable expression in our data were ignored. The sets of genes defining the modules used in our analysis are listed in the Supplemental table N.

### Statistics

The statistical tools, methods and significance thresholds for each analysis are described in the Results or Methods section or in the figure legends.

## Data Availability

The raw data have been deposited with the Gene Expression Omnibus (www.ncbi.nlm.nih.gov/geo) with the GEO accession, GSE167363. Other supporting raw data are available from the corresponding author upon request. Source data are provided with this paper.

https://www.ncbi.nlm.nih.gov/geo/query/acc.cgi?acc=GSE167363

## Data availability

The raw data have been deposited with the Gene Expression Omnibus (www.ncbi.nlm.nih.gov/geo) and the GEO accession, currently underway, will be provided in the final version of the manuscript. Other supporting raw data are available from the corresponding author upon request. Source data are provided with this paper.

## Code availability

Experimental protocols and the data analysis pipeline used in our work follow the 10X Genomics and Seurat official websites. The analysis steps, functions and parameters used are described in detail in the Methods section. Custom scripts for analyzing data are available upon reasonable request. Source data are provided with this paper.

## Acknowledgements

This research was supported by the UCR School of Medicine (to AG and MGN), the Dean Innovation Fund (to JB and MGN), and the National Institutes of Health (NIAID, R21AI37830 and R01AI153195 to MGN). The data from this study was generated at the UC San Diego IGM Genomics Center utilizing an Illumina NovaSeq 6000 that was purchased with funding from a National Institutes of Health SIG grant (#S10 OD026929). We thank Dr. Hashini Batugedara, Dr. Luqman Nasouf, Dr. Aarti Mittal, Mr. Joseph Miller, Mr. Sang Woo, the RUHS-MC sepsis response team and the Riverside Free Clinic for assistance with participant enrollment, blood collection and blood processing, and Dr. Karine Le Roch for access to the 10X equipment.

## Authorship

MGN, AG, XQ, JL, JB and WK conceptualized the study. JL, XQ, MGN and AG developed the methodology. MGN, AG, XQ, JL, JB performed the investigation. MGN, AG, XQ, JL, LJ provided the formal analysis. XQ, AG, LJ, MGN wrote the article. MGN and AG supervised the study.

