## Supplementary materials for "Dynamic changes in human single cell transcriptional signatures during fatal sepsis"

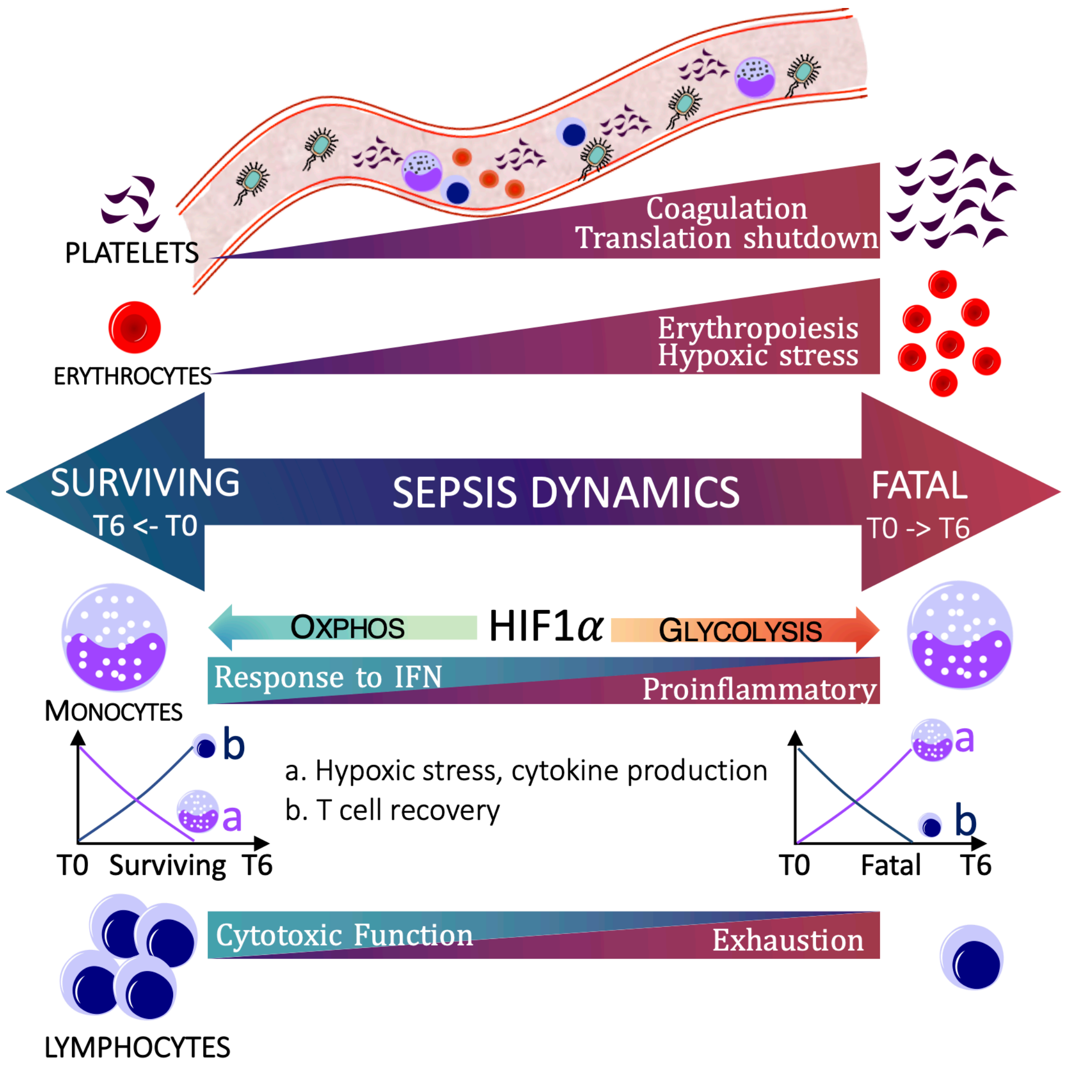


**Graphical abstract. Dynamic peripheral immune changes in sepsis survival and fatal outcomes**

### Supplementary materials

#### Supplementary methods

##### The alternative bioinformatics pipelines

We applied three alternative bioinformatics pipelines (A,B and C) as illustrated in Figure 2A. The consensus-based Method A is described in the main Methods section and its results were used for all the downstream analyses in this paper. The two other methods are described below.

**Method *B***: The MAGIC algorithm [1] was used for imputation of the normalized matrix expression values. The classification of the matrix after imputation gave results consistent with Method *A* for most of the cell types. Additionally, it identified neutrophils (1.22% in S samples, 0.16% in the NS, and 0.01% in HCs).

**Method *C***: In the profiling of single-cell transcriptomics of the immune system, the datasets are usually processed using a workflow that consists of unsupervised clustering and annotation of cell types by canonical markers. However, canonical markers applied in different studies tend to result in different cell type annotations. To further validate our results from method A and method B, we applied method C by using Seurat v4 [2] to map our query PBMC datasets to multimodal references without the unsupervised clustering and cell type annotation steps.

##### Comparison of results from different methods:

The classification of the imputation matrix (method B) gave consistent results for most of the cell types. In the healthy controls, the majority of the cells were CD4+ T cells (31.19%), followed by B cells (24.48%), platelets (0.72%), and erythroid precursor cells (0.01%). In the sepsis survivor, most of the cells were CD14+ monocytes (52.44%), followed by platelets (3.39%), and erythroid precursor cells (1.13%). In the non-survivor, most of the cells were platelets (37.04%) and erythroid precursor cells (10.51%). Also, neutrophils were not detected in method A, but showed up in the MAGIC imputation-based method B, where the sepsis survivor had the highest fraction of the neutrophils (1.22%) followed by 0.16% in the non-survivor and 0.01% in heathy controls (Supplementary Figure 1A).

The results from method C were mostly consistent with methods A and B, with more refined cell types' annotation due to the in-depth categorization of the lymphocytes according to their activation status. The subtypes recognized solely by method *C* included: CD4+ T cells with cytotoxic activity (CD4 CTL), Naive CD4+ T cells (CD4 Naïve), proliferating CD4+ T cells (CD4 Proliferating), CD4+ central memory T cells (CD4 TCM), CD4+ effector memory T cells (CD4 TEM). According to method *C,* these subtypes of CD4+ cells comprised 20.94% of all cells in healthy controls. This fraction was much lower in S samples (9.13%), and even lower in NS (6.5%). The similar trend was observed for B cells, whose subtypes recognized by method C included B intermediate, B memory and B naïve cells. In HCs, S and NS, the fraction of B cells was 25.13%, 9.53%, and 4.09%, respectively. According to method C, platelets, comprised only 1.87% of cells in HC but they were expanded to 7.39% in S, and dramatically expanded in NS (41.25%). The fractions of erythroid precursor cells in HCs, S and NS were 0.04%, 3.1%, and 19.43%, respectively. The fractions of cell types in different conditions as classified by method *C* are shown in the Supplementary Figure 1B.

##### Supplementary Materials References


1. van Dijk, D., et al., *Recovering Gene Interactions from Single-Cell Data Using Data Diffusion.* Cell, 2018. **174**(3): p. 716-729 e27.
2. Hao, Y., et al., *Integrated analysis of multimodal single-cell data.* bioRxiv, 2020.
3. Supek, F., et al., *REVIGO summarizes and visualizes long lists of gene ontology terms.* PLoS One, 2011. **6**(7): p. e21800.

### Supplementary figures

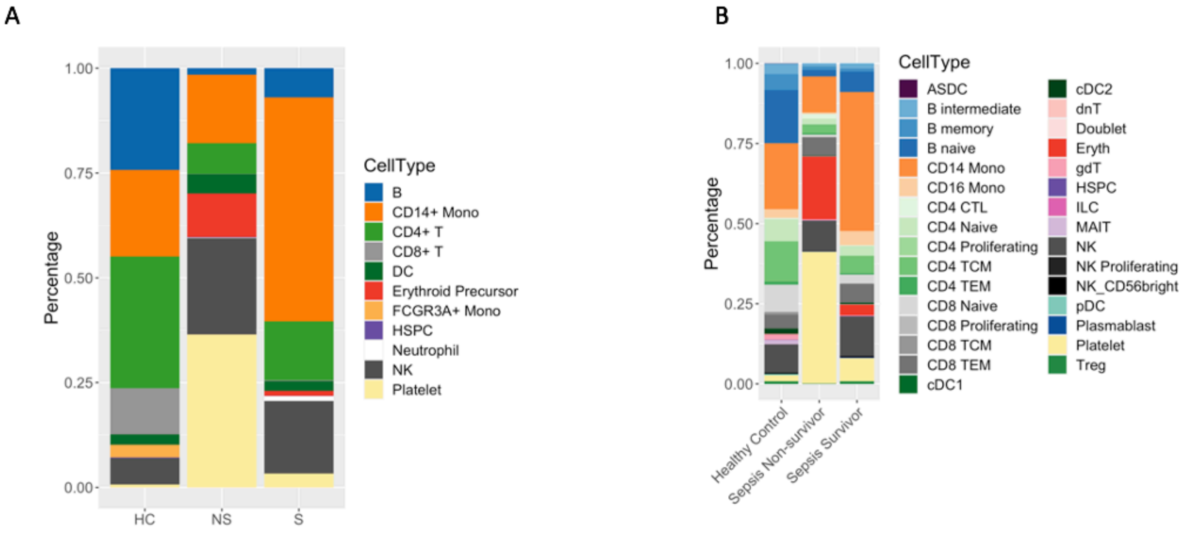


**Supplementary Figure 1. Single-cell transcriptional profiling of PBMC from healthy controls (HC) and patients with sepsis (S - survivor, NS – non-survivor) calculated with the two alternative validation methods. A,B)** Distributions of cell-types in each condition calculated with: **A)** MAGIC imputation (method *B*) **B)** mapping to the PBMC reference with Seurat v.4 (method *C*)

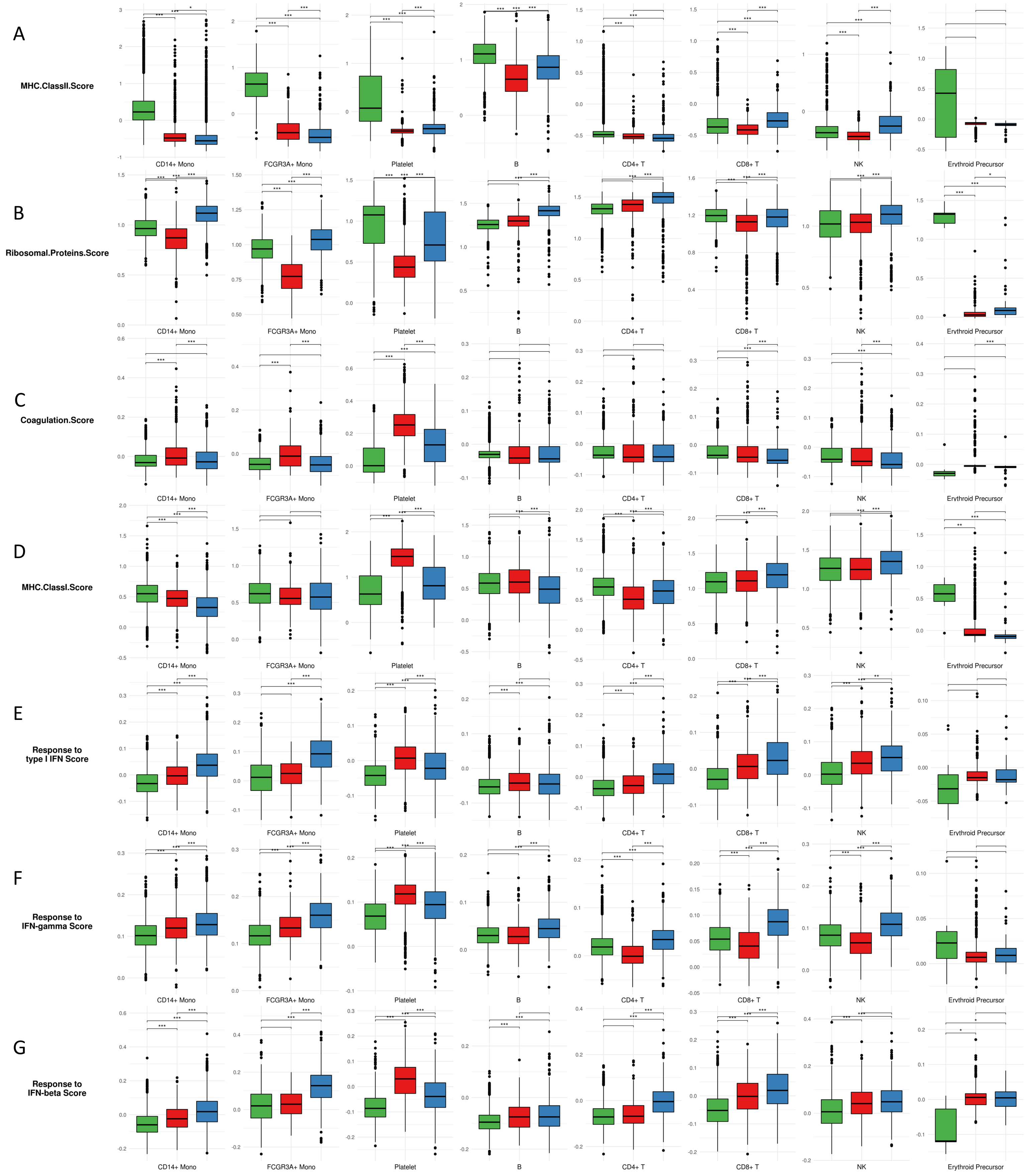


**Supplementary Figure 2. Comparisons of module scores across three conditions.**
The included modules contain genes related to: **A)** MHC Class II, **B)** Ribosomal proteins, **C)** Coagulation, **D)** MHC Class I, **E)** Response to type I IFN, **F)** Response to IFN-gamma, **G)** Response to IFN-beta. The differences in scoreS associated with Bonferroni-adjusted p-values below 0.01, 0.001, and 0.0001 are indicated as *, **, and ***, respectively. The significance analysis was performed using two-sample t-tests.

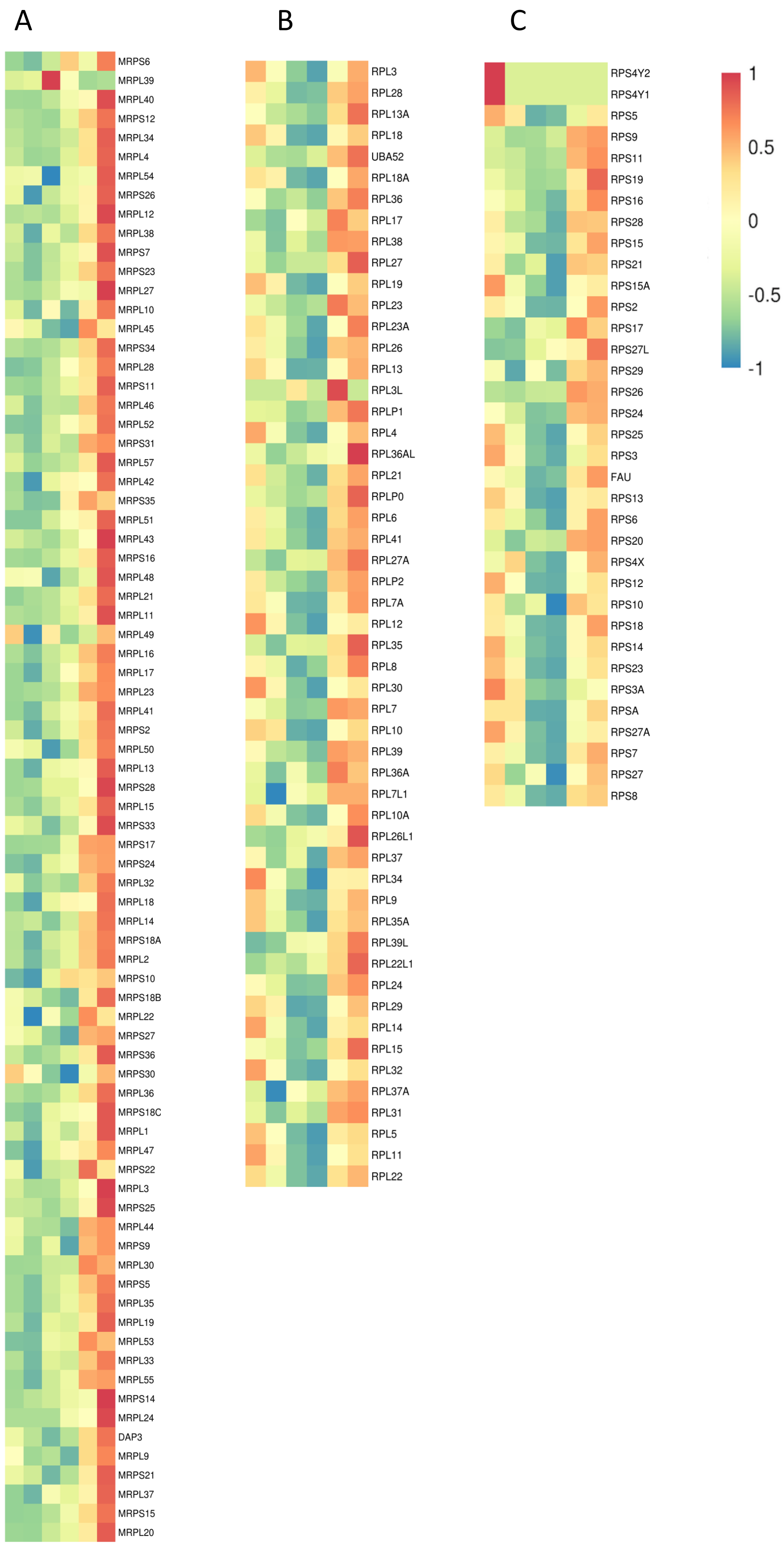


**Supplementary Figure 3. Expression of genes encoding ribosomal proteins in all samples.**
**A)** Mitochondrial ribosomal proteins. **B)** L ribosomal proteins (RPL). **C)** S ribosomal proteins (RPS). Heatmap coloring represents z-scored log-normalized mean gene expression counts averaged across all cells from a given sample.

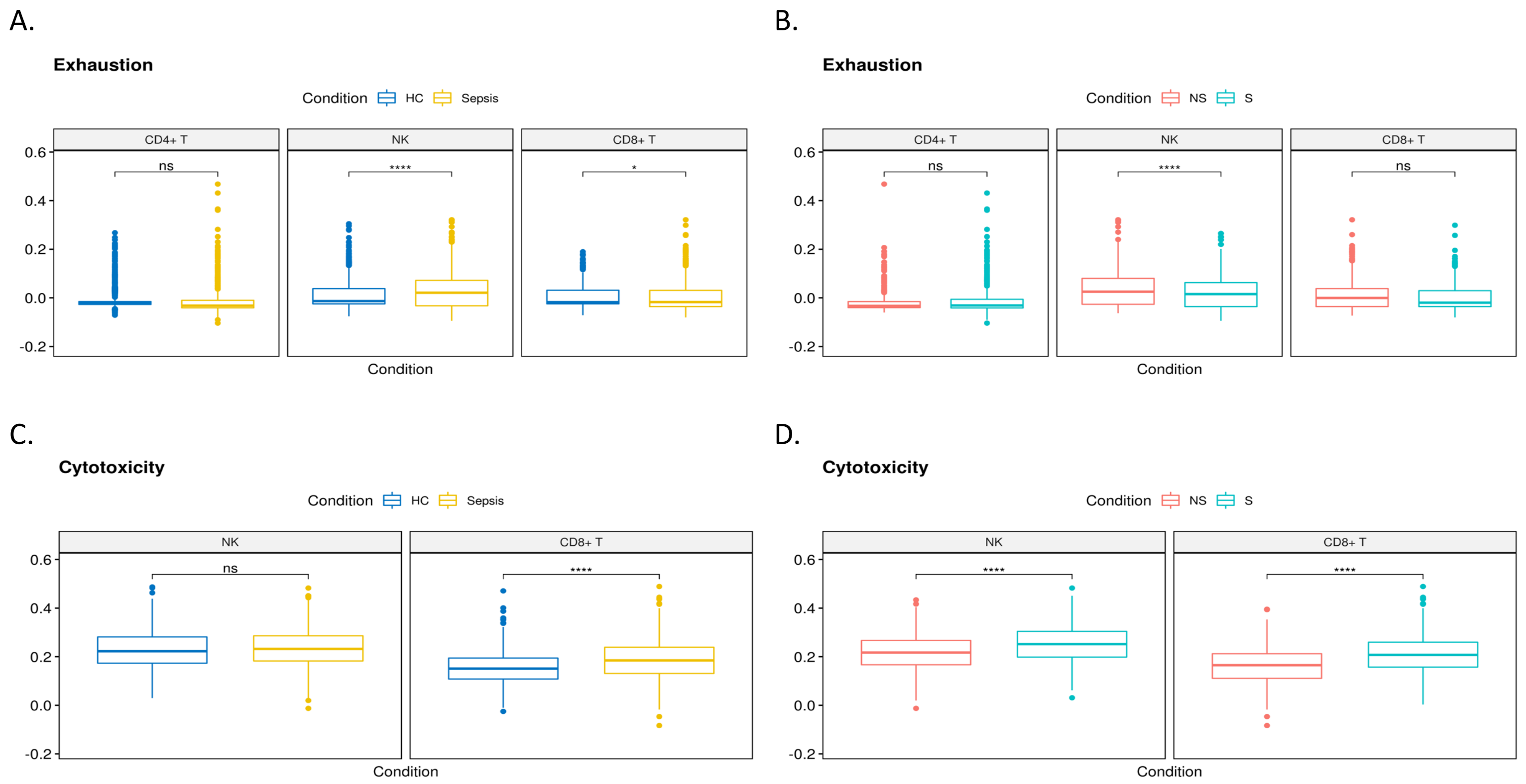


**Supplementary Figure 4. Assessment of lymphocyte activity changes between conditions.**
**A, B)** Expression score of gene module representing exhaustion: **A)** Comparison of healthy controls (HCs) vs. sepsis patients, **B)** non-survivor vs. survivor.
**C, D)** Expression score of gene module representing cytotoxicity: **C)** Comparison of healthy controls (HCs) vs. sepsis patients, **D)** non-survivor vs. survivor.
The differences with Bonferroni-adjusted p-values below 0.01, 0.001, and 0.0001 are indicated as *, **, and ***, respectively. The significance analysis was performed using two-sample t-tests.

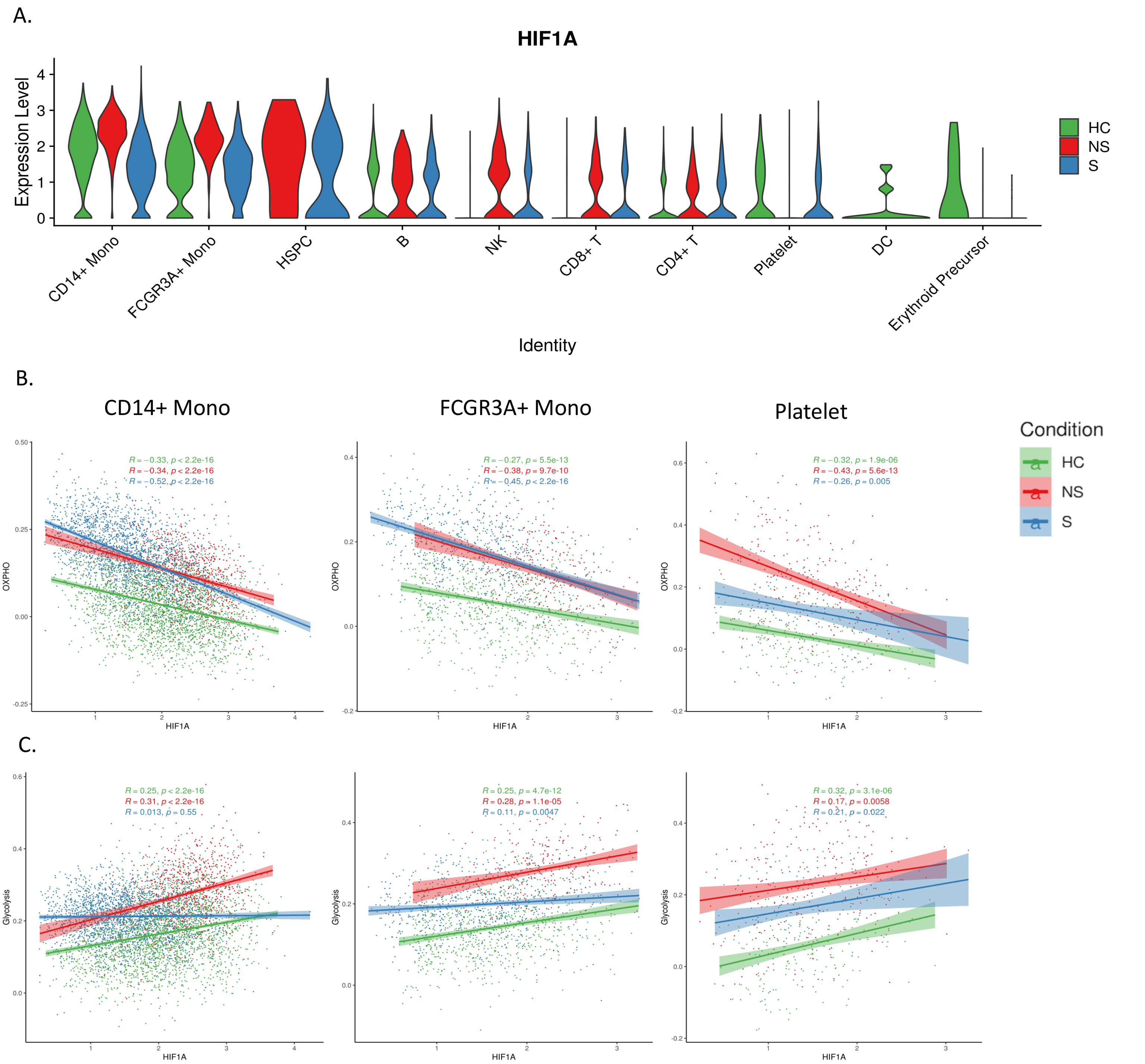


**Supplementary Figure 5. HIF1A expression and its correlation with the metabolic functions.**
**A)** The expression of the HIF1A gene in different cell types across three conditions.
**B, C)** The correlations between the HIF1A expression and: **B)** module score for oxidative phosphorylation (OXPHOS) and **C)** glycolysis module score, in: CD14+ monocytes, FCGR3A+ monocytes and platelets in each condition. R-values from Pearson's correlation, exact two-sided p-values and the 95% confidence intervals are shown on each graph. Each dot represents a single cell. Only cells with HIF1A expression ≠ 0 were included in the analysis. Green, red and blue points represent cells from HC, NS and S samples, respectively.

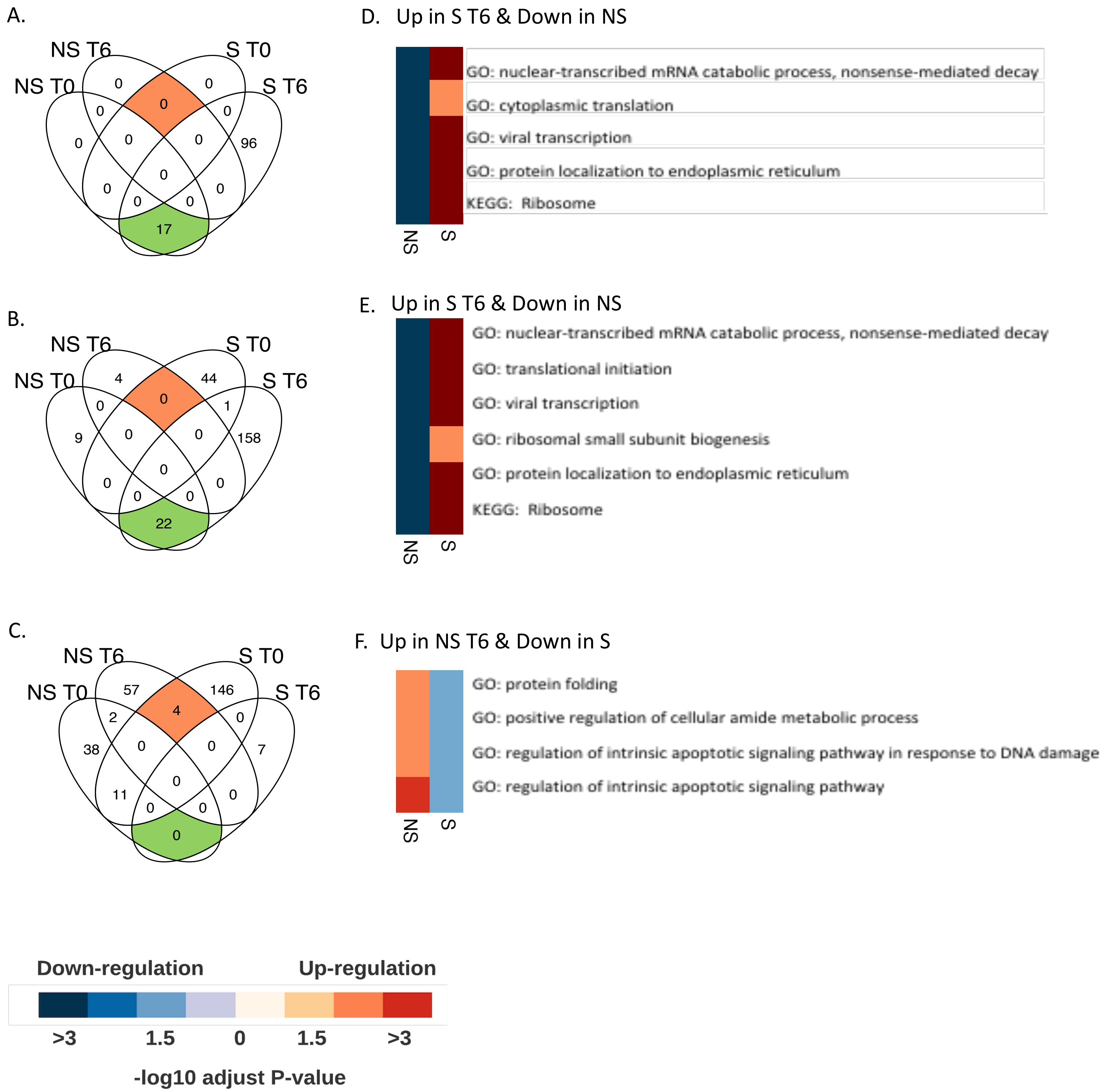


**Supplementary Figure 6. Comparison of temporal pathway changes in sepsis non-survivor and survivor.** Comparison of temporal changes in pathway expression between sepsis non-survivor and survivor. Venn diagrams describing temporal changes in pathways from each sepsis patients in **A)** CD8+ T cells, **B)** NK-cells, **C)** Platelets. Set of pathways increasing in NS (up-regulated in NS T6 as compared to NS T0) but decreasing in S (down-regulated in S T6 as compared to S T0) is colored in orange. Set of pathways decreasing from NS T0 to T6 but increasing from S T0 to T6 is colored in green.

**D,E)** Heatmap illustration of temporal changes of pathways which are increasing in S but decreasing in NS (indicated by green coloring in Venn diagrams) in **D)** CD8+ T cells and **E)** in NK cells. **F)** Heatmap illustration of pathways which are increasing in NS but decreasing in S (marked with orange coloring in Venn diagrams) in platelets. The sets of overlapping GO terms were reduced to representative ones using Revigo [3] (the cutoffs were: more than 10 overlapping GO terms and similarity > 0.4).

SUPPLEMENTARY TABLE 1

| MHC class II | Ribosomal proteins | Coagulation | MHC class I | Response to type I IFN | Response to interferon-gamma | Response to interferon-beta | Exhaustion | Cytotoxicity | Cytokine activity | OXPHOS | Glycolysis |
|---|---|---|---|---|---|---|---|---|---|---|---|
| CD74 | DAP3 | F12 | HLA-A | ADAR | ACTG1 | AIM2 | PDCD1 | AGER | AIMP1 | ACTN3 | ALDOA |
| HLA-DMA | FAU | F13A1 | HLA-B | CACTIN | ACTR2 | BST2 | HAVCR2 | B2M | AREG | AK2 | ALDOC |
| HLA-DMB | MRPL1 | F5 | HLA-C | CDC37 | ACTR3 | CDC34 | LAG3 | CTSC | BMP1 | ATP5F1C | ARNT |
| HLA-DOA | MRPL1 | F7 | HLA-E | CNOT7 | ARG1 | DDX41 | CD244 | CTSH | BMP2 | ATP5F1D | ENO1 |
| HLA-DOB | MRPL10 | F8 | HLA-F | DCST1 | BST2 | GBP2 | ENTPD1 | EMP2 | BMP3 | ATP7A | ENO2 |
| HLA-DPA1 | MRPL10 | GGCX | | FADD | CAPG | GBP3 | CD38 | FADD | BMP4 | CHCHD10 | ENO3 |
| HLA-DPB1 | MRPL11 | GP1BA | | IFITM1 | CASP1 | HTRA2 | CD101 | GZMB | BMP5 | COQ7 | ENTPD5 |
| HLA-DQA1 | MRPL11 | KLKB1 | | IFITM2 | CCL2 | IFIT1 | TIGIT | GZMM | BMP6 | COQ9 | GAPDH |
| HLA-DQA2 | MRPL12 | KNG1 | | IFITM3 | CCL3 | IFIT3 | CTLA4 | HPRT | BMP7 | COX10 | GAPDHS |
| HLA-DQB1 | MRPL12 | LMAN1 | | IFNAR1 | CCL4 | IFITM1 | TOX | HSPA8 | BMP8A | COX15 | GCK |
| HLA-DQB2 | MRPL13 | MCFD2 | | IFNAR2 | CCL5 | IFITM2 | NR4A1 | IL7R | BMP8B | COX4I1 | GPI |
| HLA-DRA | MRPL13 | PLG | | IKBKE | CCL7 | IFITM3 | IRF4 | IL12A | BMP10 | COX5A | HIF1A |
| HLA-DRB1 | MRPL14 | SERPINE1 | | IRAK1 | CCL20 | IFNAR2 | | IL23A | BMP15 | COX8A | HK1 |
| HLA-DRB5 | MRPL14 | SERPINF2 | | IRF3 | CCL22 | IKBKE | | KLRC1 | C1QTNF4 | CYCS | HK2 |
| | MRPL15 | VKORC1 | | IRF7 | CCL26 | IRF1 | | KLRD1 | CCL1 | DLD | HK3 |
| | MRPL15 | VWF | | ISG15 | CD40 | IRGM2 | | MR1 | CCL2 | DNAJC15 | HTR2A |
| | MRPL16 | | | LSM14A | CD47 | NDUFA13 | | NECTIN2 | CCL3 | FXN | INSR |
| | MRPL16 | | | MAVS | CD74 | PLSCR1 | | P2RX7 | CCL4 | GADD45GIP1 | MYC |
| | MRPL17 | | | METTL3 | CDC37 | PNPT1 | | PNP | CCL5 | LEXM | P2RX7 |
| | MRPL17 | | | MUL1 | CDC42 | STAT1 | | PPP3CB | CCL7 | MECP2 | PFKFB2 |
| | MRPL18 | | | MYD88 | CDC42EP2 | TRIM6 | | PRF1 | CCL8 | MLXIPL | PFKFB3 |
| | MRPL18 | | | NLRC5 | CDC42EP4 | UBE2G2 | | PTPRC | CCL11 | MSH2 | PFKFB4 |
| | MRPL19 | | | OAS2 | CIITA | UBE2K | | RAB27A | CCL17 | NDUFA1 | PFKL |
| | MRPL19 | | | PTPN2 | CITED1 | XAF1 | | RIPK3 | CCL19 | NDUFA10 | PFKM |
| | MRPL2 | | | SAMHD1 | CXCL16 | | | SERPINB9 | CCL20 | NDUFA2 | PFKP |
| | MRPL2 | | | SETD2 | CYP27B1 | | | STX7 | CCL22 | NDUFA3 | PGAM1 |
| | MRPL20 | | | SHMT2 | DAPK1 | | | STX11 | CCL24 | NDUFA4 | PGK1 |
| | MRPL20 | | | SMPD1 | DAPK3 | | | TAP2 | CCL25 | NDUFA5 | PPP2R5D |
| | MRPL21 | | | STAT1 | DNAJA3 | | | XCL1 | CCL26 | NDUFA6 | PRKAA1 |
| | MRPL21 | | | STAT2 | EPRS | | | | CCL28 | NDUFA7 | TPI1 |
| | MRPL22 | | | TBK1 | EVL | | | | CD40LG | NDUFA8 | |
| | MRPL22 | | | TRIM6 | FLNB | | | | CD70 | NDUFA9 | |
| | MRPL23 | | | TRIM56 | GAPDH | | | | CER1 | NDUFAB1 | |
| | MRPL23 | | | TTLL12 | GBP2 | | | | CKLF | NDUFAF1 | |
| | MRPL24 | | | UBE2K | GBP3 | | | | CLCF1 | NDUFB1 | |
| | MRPL24 | | | WNT5A | GBP4 | | | | CMTM3 | NDUFB10 | |
| | MRPL27 | | | YTHDF2 | GBP5 | | | | CMTM5 | NDUFB2 | |
| | MRPL27 | | | YTHDF3 | GBP7 | | | | CMTM7 | NDUFB3 | |
| | MRPL28 | | | ZBP1 | GCH1 | | | | CMTM8 | NDUFB4 | |
| | MRPL28 | | | | GSN | | | | CNTF | NDUFB5 | |
| | MRPL3 | | | | IFITM1 | | | | CRLF1 | NDUFB6 | |
| | MRPL3 | | | | IFITM2 | | | | CRLF2 | NDUFB7 | |
| | MRPL30 | | | | IFITM3 | | | | CSF1 | NDUFB8 | |
| | MRPL30 | | | | IFNG | | | | CSF2 | NDUFB9 | |
| | MRPL32 | | | | IL12RB1 | | | | CSF3 | NDUFC1 | |
| | MRPL32 | | | | IL23R | | | | CTF1 | NDUFC2 | |
| | MRPL33 | | | | IRF1 | | | | CEBPZ | NDUFS1 | |
| | MRPL33 | | | | IRF8 | | | | CX3CL1 | NDUFS2 | |
| | MRPL34 | | | | IRGM | | | | CXCL1 | NDUFS3 | |
| | MRPL34 | | | | JAK2 | | | | CXCL2 | NDUFS4 | |
| | MRPL35 | | | | KIF5B | | | | CXCL3 | NDUFS5 | |
| | MRPL35 | | | | KIF16B | | | | CXCL5 | NDUFS6 | |
| | MRPL36 | | | | KYNU | | | | CXCL9 | NDUFS7 | |
| | MRPL36 | | | | MED1 | | | | CXCL10 | NDUFS8 | |
| | MRPL37 | | | | MEFV | | | | CXCL11 | NDUFV1 | |
| | MRPL37 | | | | MRC1 | | | | CXCL12 | NDUFV2 | |
| | MRPL38 | | | | MYO1C | | | | CXCL13 | NDUFV3 | |
| | MRPL38 | | | | MYO18A | | | | CXCL14 | NIPSNAP2 | |
| | MRPL39 | | | | NLRC5 | | | | CXCL16 | PARK7 | |
| | MRPL39 | | | | NMI | | | | CXCL17 | PINK1 | |

| MHC class II | Ribosomal proteins | Coagulation | MHC class I | Response to type I IFN | Response to interferon-gamma | Response to interferon-beta | Exhaustion | Cytotoxicity | Cytokine activity | OXPHOS | Glycolysis |
|---|---|---|---|---|---|---|---|---|---|---|---|
| | MRPL4 | | | | PARP9 | | | | EBI3 | PMPCB | |
| | MRPL4 | | | | PARP14 | | | | EDN1 | PPIF | |
| | MRPL40 | | | | PDE12 | | | | EPO | SDHAF2 | |
| | MRPL40 | | | | PPARG | | | | FAM3B | SDHC | |
| | MRPL41 | | | | PTPN2 | | | | FASLG | SLC25A23 | |
| | MRPL41 | | | | RAB11FIP5 | | | | FGF2 | SLC25A33 | |
| | MRPL42 | | | | RAB12 | | | | GDF1 | SNCA | |
| | MRPL42 | | | | RAB20 | | | | GDF2 | SURF1 | |
| | MRPL43 | | | | RAB43 | | | | GDF3 | TAZ | |
| | MRPL43 | | | | RPL13A | | | | GDF5 | UQCR10 | |
| | MRPL44 | | | | RPS6KB1 | | | | GDF6 | UQCRB | |
| | MRPL44 | | | | SIRPA | | | | GDF7 | UQCRC1 | |
| | MRPL45 | | | | SLC11A1 | | | | GDF9 | UQCRC2 | |
| | MRPL45 | | | | SLC26A6 | | | | GDF10 | UQCRH | |
| | MRPL46 | | | | SNCA | | | | GDF11 | UQCRHL | |
| | MRPL46 | | | | SOCS1 | | | | GDF15 | VCP | |
| | MRPL47 | | | | STAT1 | | | | PIGQ | | |
| | MRPL47 | | | | STX4 | | | | GREM1 | | |
| | MRPL48 | | | | STX8 | | | | GREM2 | | |
| | MRPL48 | | | | STX11 | | | | GRN | | |
| | MRPL49 | | | | STXBP1 | | | | HMGB1 | | |
| | MRPL49 | | | | STXBP2 | | | | IFNA1 | | |
| | MRPL50 | | | | STXBP3 | | | | IFNA2 | | |
| | MRPL50 | | | | STXBP4 | | | | IFNA4 | | |
| | MRPL51 | | | | SYNCRIP | | | | IFNA5 | | |
| | MRPL51 | | | | TLR2 | | | | IFNA6 | | |
| | MRPL52 | | | | TLR4 | | | | IFNA7 | | |
| | MRPL52 | | | | TRIM21 | | | | IFNA13 | | |
| | MRPL53 | | | | TXK | | | | IFNA14 | | |
| | MRPL53 | | | | VAMP3 | | | | IFNA16 | | |
| | MRPL54 | | | | VAMP4 | | | | IFNB1 | | |
| | MRPL54 | | | | VAMP8 | | | | IFNE | | |
| | MRPL55 | | | | VIM | | | | IFNG | | |
| | MRPL55 | | | | VPS26B | | | | IFNK | | |
| | MRPL57 | | | | WAS | | | | IFNL2 | | |
| | MRPL57 | | | | XCL1 | | | | IFNL3 | | |
| | MRPL58 | | | | ZYX | | | | IL1A | | |
| | MRPL9 | | | | | | | | IL1B | | |
| | MRPS10 | | | | | | | | IL1F10 | | |
| | MRPS10 | | | | | | | | IL1RN | | |
| | MRPS11 | | | | | | | | IL2 | | |
| | MRPS11 | | | | | | | | IL3 | | |
| | MRPS12 | | | | | | | | IL4 | | |
| | MRPS12 | | | | | | | | IL5 | | |
| | MRPS14 | | | | | | | | IL6 | | |
| | MRPS14 | | | | | | | | IL7 | | |
| | MRPS15 | | | | | | | | IL9 | | |
| | MRPS15 | | | | | | | | IL10 | | |
| | MRPS16 | | | | | | | | IL11 | | |
| | MRPS16 | | | | | | | | IL12A | | |
| | MRPS17 | | | | | | | | IL12B | | |
| | MRPS17 | | | | | | | | IL13 | | |
| | MRPS18A | | | | | | | | IL15 | | |
| | MRPS18A | | | | | | | | IL16 | | |
| | MRPS18B | | | | | | | | IL17A | | |
| | MRPS18B | | | | | | | | IL17B | | |
| | MRPS18C | | | | | | | | IL17C | | |
| | MRPS18C | | | | | | | | IL17D | | |
| | MRPS2 | | | | | | | | IL17F | | |
| | MRPS2 | | | | | | | | IL18 | | |

SUPPLEMENTARY TABLE 1

| MHC class II | Ribosomal proteins | Coagulation | MHC class I | Response to type I IFN | Response to interferon-gamma | Response to interferon-beta | Exhaustion | Cytotoxicity | Cytokine activity | OXPHOS | Glycolysis |
|---|---|---|---|---|---|---|---|---|---|---|---|
| | MRPS21 | | | | | | | | IL19 | | |
| | MRPS21 | | | | | | | | IL20 | | |
| | MRPS22 | | | | | | | | IL21 | | |
| | MRPS22 | | | | | | | | IL22 | | |
| | MRPS23 | | | | | | | | IL23A | | |
| | MRPS23 | | | | | | | | IL24 | | |
| | MRPS24 | | | | | | | | IL25 | | |
| | MRPS24 | | | | | | | | IL27 | | |
| | MRPS25 | | | | | | | | IL31 | | |
| | MRPS25 | | | | | | | | IL33 | | |
| | MRPS26 | | | | | | | | IL34 | | |
| | MRPS26 | | | | | | | | IL36A | | |
| | MRPS27 | | | | | | | | IL36B | | |
| | MRPS27 | | | | | | | | IL36G | | |
| | MRPS28 | | | | | | | | IL36RN | | |
| | MRPS28 | | | | | | | | INHA | | |
| | MRPS30 | | | | | | | | INHBA | | |
| | MRPS30 | | | | | | | | INHBB | | |
| | MRPS31 | | | | | | | | INHBC | | |
| | MRPS31 | | | | | | | | INHBE | | |
| | MRPS33 | | | | | | | | KITLG | | |
| | MRPS33 | | | | | | | | LEFTY1 | | |
| | MRPS34 | | | | | | | | LEFTY2 | | |
| | MRPS34 | | | | | | | | LIF | | |
| | MRPS35 | | | | | | | | LTA | | |
| | MRPS35 | | | | | | | | LTB | | |
| | MRPS36 | | | | | | | | MIF | | |
| | MRPS36 | | | | | | | | MSMP | | |
| | MRPS5 | | | | | | | | MSTN | | |
| | MRPS6 | | | | | | | | NAMPT | | |
| | MRPS7 | | | | | | | | NDP | | |
| | MRPS9 | | | | | | | | NODAL | | |
| | RPL10 | | | | | | | | OSM | | |
| | RPL10ARPL11 | | | | | | | | PF4 | | |
| | RPL12 | | | | | | | | PGLYRP1 | | |
| | RPL13 | | | | | | | | PPBP | | |
| | RPL13A | | | | | | | | SCG2 | | |
| | RPL14 | | | | | | | | SCGB3A1 | | |
| | RPL15 | | | | | | | | SLURP1 | | |
| | RPL17 | | | | | | | | SPP1 | | |
| | RPL18 | | | | | | | | TGFB1 | | |
| | RPL18A | | | | | | | | TGFB2 | | |
| | RPL19 | | | | | | | | TGFB3 | | |
| | RPL21 | | | | | | | | THPO | | |
| | RPL22 | | | | | | | | TIMP1 | | |
| | RPL22L1 | | | | | | | | TNF | | |
| | RPL23 | | | | | | | | TNFSF4 | | |
| | RPL23A | | | | | | | | TNFSF8 | | |
| | RPL24 | | | | | | | | TNFSF9 | | |
| | RPL26 | | | | | | | | TNFSF10 | | |
| | RPL26L1 | | | | | | | | TNFSF11 | | |
| | RPL27 | | | | | | | | TNFSF12 | | |
| | RPL27A | | | | | | | | TNFSF13 | | |
| | RPL28 | | | | | | | | TNFSF13B | | |
| | RPL29 | | | | | | | | TNFSF14 | | |
| | RPL30 | | | | | | | | TNFSF15 | | |
| | RPL31 | | | | | | | | TNFSF18 | | |
| | RPL32 | | | | | | | | TSLP | | |
| | RPL34 | | | | | | | | VEGFA | | |
| | RPL35 | | | | | | | | WNT1 | | |

| MHC class II | Ribosomal proteins | Coagulation | MHC class I | Response to type I IFN | Response to interferon-gamma | Response to interferon-beta | Exhaustion | Cytotoxicity | Cytokine activity | OXPHOS | Glycolysis |
|---|---|---|---|---|---|---|---|---|---|---|---|
| | RPL35A | | | | | | | | WNT2 | | |
| | RPL36 | | | | | | | | WNT2B | | |
| | RPL36A | | | | | | | | WNT3 | | |
| | RPL36AL | | | | | | | | WNT3A | | |
| | RPL37 | | | | | | | | WNT4 | | |
| | RPL37A | | | | | | | | WNT5A | | |
| | RPL38 | | | | | | | | WNT5B | | |
| | RPL39 | | | | | | | | WNT6 | | |
| | RPL39L | | | | | | | | WNT7A | | |
| | RPL3RPL4 | | | | | | | | WNT7B | | |
| | RPL41 | | | | | | | | WNT8A | | |
| | RPL5 | | | | | | | | WNT8B | | |
| | RPL6 | | | | | | | | WNT9A | | |
| | RPL7 | | | | | | | | WNT9B | | |
| | RPL7A | | | | | | | | WNT10A | | |
| | RPL7L1 | | | | | | | | WNT10B | | |
| | RPL8 | | | | | | | | WNT11 | | |
| | RPL9 | | | | | | | | WNT16 | | |
| | RPLP0 | | | | | | | | XCL1 | | |
| | RPLP1 | | | | | | | | | | |
| | RPLP2 | | | | | | | | | | |
| | RPS10 | | | | | | | | | | |
| | RPS11 | | | | | | | | | | |
| | RPS12 | | | | | | | | | | |
| | RPS13 | | | | | | | | | | |
| | RPS14 | | | | | | | | | | |
| | RPS15 | | | | | | | | | | |
| | RPS15A | | | | | | | | | | |
| | RPS16 | | | | | | | | | | |
| | RPS17 | | | | | | | | | | |
| | RPS18 | | | | | | | | | | |
| | RPS19 | | | | | | | | | | |
| | RPS2 | | | | | | | | | | |
| | RPS20 | | | | | | | | | | |
| | RPS21 | | | | | | | | | | |
| | RPS23 | | | | | | | | | | |
| | RPS24 | | | | | | | | | | |
| | RPS25 | | | | | | | | | | |
| | RPS26 | | | | | | | | | | |
| | RPS27 | | | | | | | | | | |
| | RPS27A | | | | | | | | | | |
| | RPS27L | | | | | | | | | | |
| | RPS28 | | | | | | | | | | |
| | RPS29 | | | | | | | | | | |
| | RPS3 | | | | | | | | | | |
| | RPS3A | | | | | | | | | | |
| | RPS4X | | | | | | | | | | |
| | RPS4Y1 | | | | | | | | | | |
| | RPS4Y2 | | | | | | | | | | |
| | RPS5 | | | | | | | | | | |
| | RPS6 | | | | | | | | | | |
| | RPS7 | | | | | | | | | | |
| | RPS8 | | | | | | | | | | |
| | RPS9 | | | | | | | | | | |
| | RPSA | | | | | | | | | | |
| | UBA52 | | | | | | | | | | |
